# Deleterious mitochondrial heteroplasmy drives high-risk clonal hematopoiesis and hematological malignancy

**DOI:** 10.64898/2026.06.22.26356227

**Authors:** Dan E. Arking, Tryggvi McDonald, Wen Shi, Daniela Puiu, Jeremy V. Arking, Sergiu Pasca, Yun Soo Hong, Lukasz Gondek

**Author notes:** Corresponding Author: Dan E. Arking, Johns Hopkins University School of Medicine 733 N. Broadway Street, MRB-459 Baltimore, MD 21205, Email of corresponding author.

## Abstract

Mitochondrial DNA (mtDNA) heteroplasmy, the coexistence of multiple mtDNA variants within cells, accumulates with age and is associated with hematological malignancies and mortality. However, whether predicted deleterious heteroplasmies causally contribute to cancer or merely represent passenger mutations remains unresolved. Here, leveraging ∼36,000 first-degree relative pairs from the UK Biobank and All of Us Research Program cohorts, we deconvolute overall heteroplasmy metrics into those that are shared across family members (representing inherited variants) and those that are not (representing de novo variants) to establish a Mendelian randomization framework for assessing causality. We show that shared heteroplasmies exhibit strong purifying selection, with reduced predicted deleteriousness compared to not shared variants, and that 90% of an individual’s deleterious heteroplasmy burden is somatically acquired. Critically, shared deleterious heteroplasmy burden, fixed at conception and thus temporally upstream of potential confounders, is significantly associated with hematological malignancies (RR=2.81, 95% CI 1.29-6.13), with effect sizes concordant with the not shared heteroplasmy burden. Furthermore, shared deleterious heteroplasmy specifically associates with high-risk clonal hematopoiesis of indeterminate potential (CHIP), particularly spliceosome mutations, suggesting mitochondrial dysfunction promotes clonal expansion of specific CHIP subtypes. Finally, we identify ultra-rare individual mtDNA variants associated with hematological malignancies, a hallmark of driver mutations. These findings establish mtDNA heteroplasmies, including inherited variants, as causal contributors to hematological malignancy risk and demonstrate that most disease-relevant burden is acquired during life, identifying potential opportunities for prevention and therapeutic intervention in individuals at elevated risk for hematological cancer, particularly of myeloid origin.

## Main

Mitochondria are central to cellular metabolism, signaling, and apoptosis, and pathogenic mitochondrial DNA (mtDNA) mutations can have profound consequences across multiple organ systems.^1^ Mitochondria are maternally inherited and maintain their own genome with 22 genes that code for key components of the translation machinery (2 ribosomal RNAs, 22 tRNAs), along with 13 genes that encode subunits of the respiratory chain (contributing to complexes I, III, IV, and V). A key feature of mtDNA is that it exists as a multi-copy genome, ranging from 10-1000s of copies per cell, allowing for a state of heteroplasmy, in which more than one mtDNA allele exists in a cell or tissue. Due, in part, to reduced efficiency of the mtDNA repair machinery, somatic mutations accumulate in mtDNA at much higher rates than in the nuclear genome.^2,3^ Indeed, we and others have demonstrated that somatic mutations accumulate non-linearly with age, and are more common in smokers.^4–6^ In a large-scale population-based cohort of individuals aged 40-70, the UK Biobank (UKB), heteroplasmy was present in ∼30% of participants.^5^

Beyond primary mitochondrial disorders, mitochondrial heteroplasmy has been associated with aging-related diseases, including chronic kidney disease,^7^ various cancers,^5,8–12^ and overall mortality.^5^ Hematological cancers, and in particular myeloid neoplasms (MN) and chronic lymphocytic leukemia (CLL), are often preceded by a pre-malignant condition, known as clonal hematopoiesis of indeterminate potential (CHIP), which is defined by the presence of cancer-associated somatic mutations in the nuclear genome in otherwise healthy individuals.^13–15^ CHIP and mitochondrial heteroplasmy often co-occur, and higher risk CHIP categories (higher variant allele fraction [VAF], multiple CHIP mutations, high-risk gene mutation) have been associated with more predicted deleterious heteroplasmy levels, suggesting a link between mtDNA heteroplasmy and CHIP.^8^ Moreover, heteroplasmy is both an independent predictor of MN and CLL risk and demonstrates a significant synergistic interaction with CHIP in predicting MN risk.^8,9^ Several lines of evidence point to a causal role for mitochondrial heteroplasmy in MN/CLL risk, including its association with MN/CLL risk in the absence of detectable CHIP and the observation that predicted deleteriousness of a heteroplasmy is associated with an increased risk even after adjusting for the number of heteroplasmies, suggesting that heteroplasmies are not simply passenger mutations.^8,9^ However, whether predicted deleterious heteroplasmies are causal contributors to malignant transformation or instead passenger events linked to clonal expansion driven by nuclear alterations remains unresolved, with important implications for prevention and therapeutic strategies.

In this work, we leverage two large biobank cohorts, the UKB and All of Us Research Program, with a combined ∼36,000 first-degree relative pairs, to deconvolute overall heteroplasmy metrics into those that are shared across family members (representing inherited variants) and those that are not (representing de novo variants). By establishing a measure of inherited heteroplasmy, we are able to use Mendelian randomization to directly assess whether mtDNA heteroplasmy is causally associated with MN/CLL risk. We also use the full cohorts, including both related and unrelated individuals, to identify highly penetrant recurrent mitochondrial mutations associated with hematological malignancies.

## Results

Among 490,270 individuals from the UKB, we identified 29,248 first degree relative pairs (180 monozytogic [MZ] twins, 6,307 Parent-Offspring, 22,761 Full Siblings). After data cleaning (see **Methods**) and limiting to 1 relative per participant, 22,154 first degree relative pairs remained for analysis (170 MZ, 3,910 Mother-Offspring, 18,074 Full Siblings) (**Supplementary Table 1**). Additionally, there were 1,698 Father-Offspring pairs available to serve as negative controls for mtDNA transmission. Heteroplasmic variants were identified in each individual using the MitoHPC software package,^4^ with heteroplasmy defined as a VAF of 0.05-0.95. For each relative pair, we assessed whether heteroplasmies were shared or not shared (**Table 1**). Across 1,698 Father-Offspring, we identified 9 shared vs. 1163 not shared variants, with a common heteroplasmy, 16093T>C (∼1% UKB population frequency), accounting for 5 false positives under the assumption that shared heteroplasmies are inherited. We therefore excluded 16093T>C, with the remaining variants demonstrating a very low false-positive rate for detection of inherited (shared) heteroplasmy (0.34%).

**Table 1:** Heteroplasmy count by relationship type.

| Relationship | Pairs (n) | Shared Heteroplasmy (n) | Not Shared Heteroplasmy (n) |
| --- | --- | --- | --- |
| MZ Twins | 170 | 45 | 26 |
| Mother-Offspring | 3,910 | 776 | 1,199 |
| Full Siblings | 18,074 | 2,573 | 7,697 |

### Impact of mitochondrial bottleneck on shared VAF correlation

Overall, we observed a high correlation between the VAF of shared alleles (**Figure 1**), with the highest correlation observed in MZ Twins (Pearson’s R=0.96), followed by Mother-Offspring (Pearson’s R=0.68), and lowest in Full Siblings (Pearson’s R=0.53), consistent with the impact of the bottleneck during oogenesis - the MZ Twins share the same bottleneck event, Mother-Offspring are separated by one bottleneck event, whereas the Full Siblings reflect the impact of two bottlenecks (one for each sibling). Consistent with prior work in large-scale human pedigrees^16^, we observed an average of 3 mtDNA units transmitted from Mother to Offspring (N_e_ = 2.97, 95% confidence interval [CI] 2.49-3.46).

**Fig. 1.**
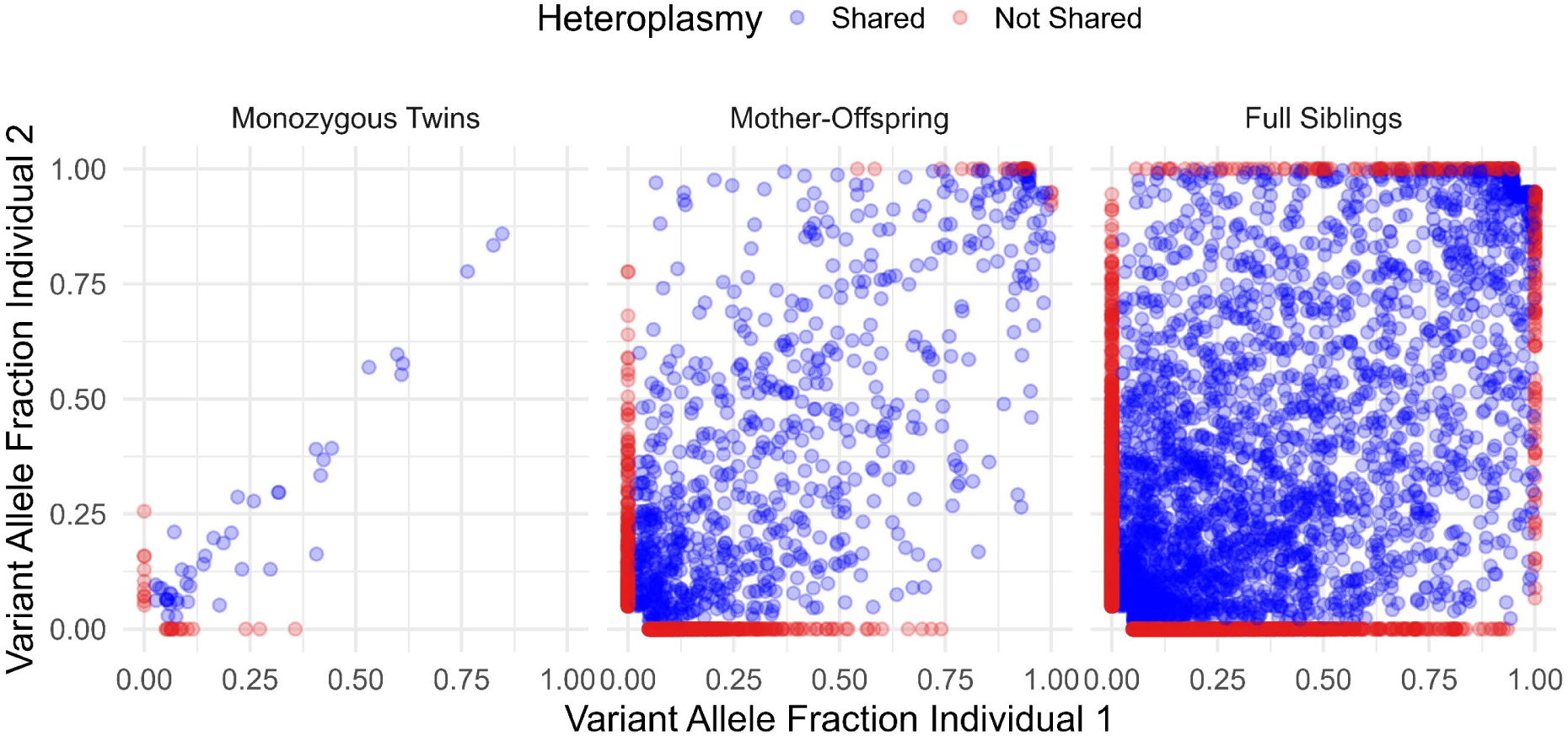
Correlation of heteroplasmy VAFs stratified by relationship type. **Left,** Monozygous Twins have 45 shared variants, Pearson’s R=0.96; **center,** Mother-Offspring have 776 shared variants, Pearson’s R=0.68; **right,** Full Siblings have 2,573 shared variants, Pearson’s R=0.53. Blue indicates shared heteroplasmies, and red indicates not shared heteroplasmies.

### mtDNA heteroplasmy characteristics for shared vs. not shared variants

Shared variants have significantly higher VAF compared to not shared variants, exhibiting a relative uniform distribution across VAF (mean VAF=0.39, median VAF=0.33), whereas most not shared variants are low VAF (mean VAF=0.15, median VAF=0.10) (**Figure 2A**), likely reflecting limited clonal expansion of not shared mutations. In contrast, not shared variants are more likely to have functional consequences (missense, P<2×10^−16^; nonsense, P<0.016) or to be found in RNA vs. D-loop regions compared to shared variants (P<8.9×10^−15^) (**Figure 2B, Supplementary Figure 1A**), and are more likely to be predicted to be deleterious based on the modified mitochondrial DNA local constraint (mMLC) score (P<2×10^−16^), which ranges from 0 to 1, with higher scores indicating more constrained, and therefore, deleterious sites^17^ (**Figure 2C, Supplementary Figure 1B**).

**Fig. 2.**
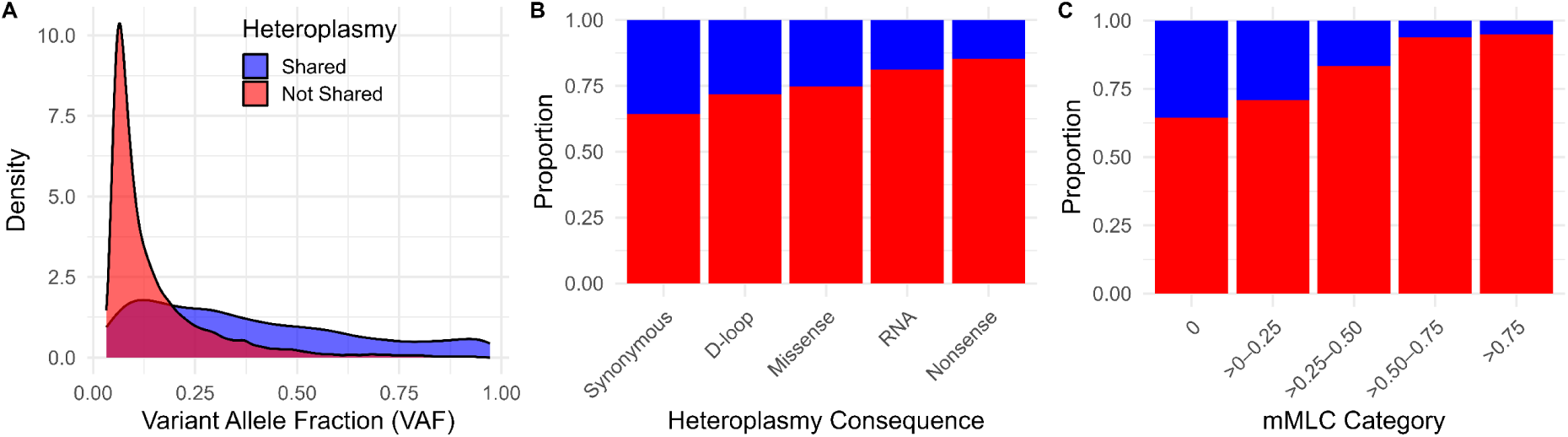
mtDNA heteroplasmy characteristics stratified by sharing status. **a,** Distribution of variant allele fraction (VAF). Shared heteroplasmy VAF is the average of the VAF for each relative pair. Not shared VAF is relative to the homoplasmy observed in the other relative. **b,** Proportion of heteroplasmies stratified by mutational consequence (synonymous, missense, nonsense) or for non-coding variation, complex (D-loop vs RNA). **c,** Proportion of heteroplasmies stratified by mMLC categories. Blue indicates shared heteroplasmies, and red indicates not shared heteroplasmies.

### Mitochondrial heteroplasmy count and the impact of maternal age at birth

For each individual, we summed the number of heteroplasmies stratified by sharing status to get shared and not shared heteroplasmy counts, as well as the overall heteroplasmy count. After adjusting for age and smoking exposure, we expected that overall heteroplasmy count would not differ by relative type (Mother, Offspring, Full Sibling, or MZ Twin). However, we observed a borderline significant association comparing a model with the relative type term to one without (P=0.053), largely driven by lower heteroplasmy count in the Offspring relative to Full Siblings (P<0.014) (**Supplementary Figure 2**). Given the study design of the UKB, which only enrolled individuals between the ages of 40-70, the maximum maternal age at birth is 30 years old, whereas the rest of the participants did not have a specific age limit, leading to, on average, younger age in Offspring compared to Full Siblings. The significant difference between Offspring and Full Siblings is still observed when limiting Full Siblings to the same age range as the Offspring (age <50 years, P<0.025). Thus, we hypothesized that maternal age at birth could be influencing heteroplasmy count. The UKB participants reported age of their parent if still alive, and we used this data to calculate the age of the parent at participant birth across 78,726 individuals with data for both maternal and paternal age in the full cohort. Both maternal (**Figure 3a**, P<5.22×10^−6^) and paternal (P<6.00×10^−5^) age were significantly associated with heteroplasmy count in separate models (**Figure 3b**). However, with both terms in the same model, only maternal age at birth was significantly associated with increased heteroplasmy count (**Figure 3b**), supporting the hypothesis that the Offspring had lower heteroplasmy count than Full Siblings due to the younger maternal age at birth. Similarly, when incorporating maternal age at birth into a regression model with the relative type, relative type was no longer associated with heteroplasmy count (P=0.53).

**Fig. 3.**
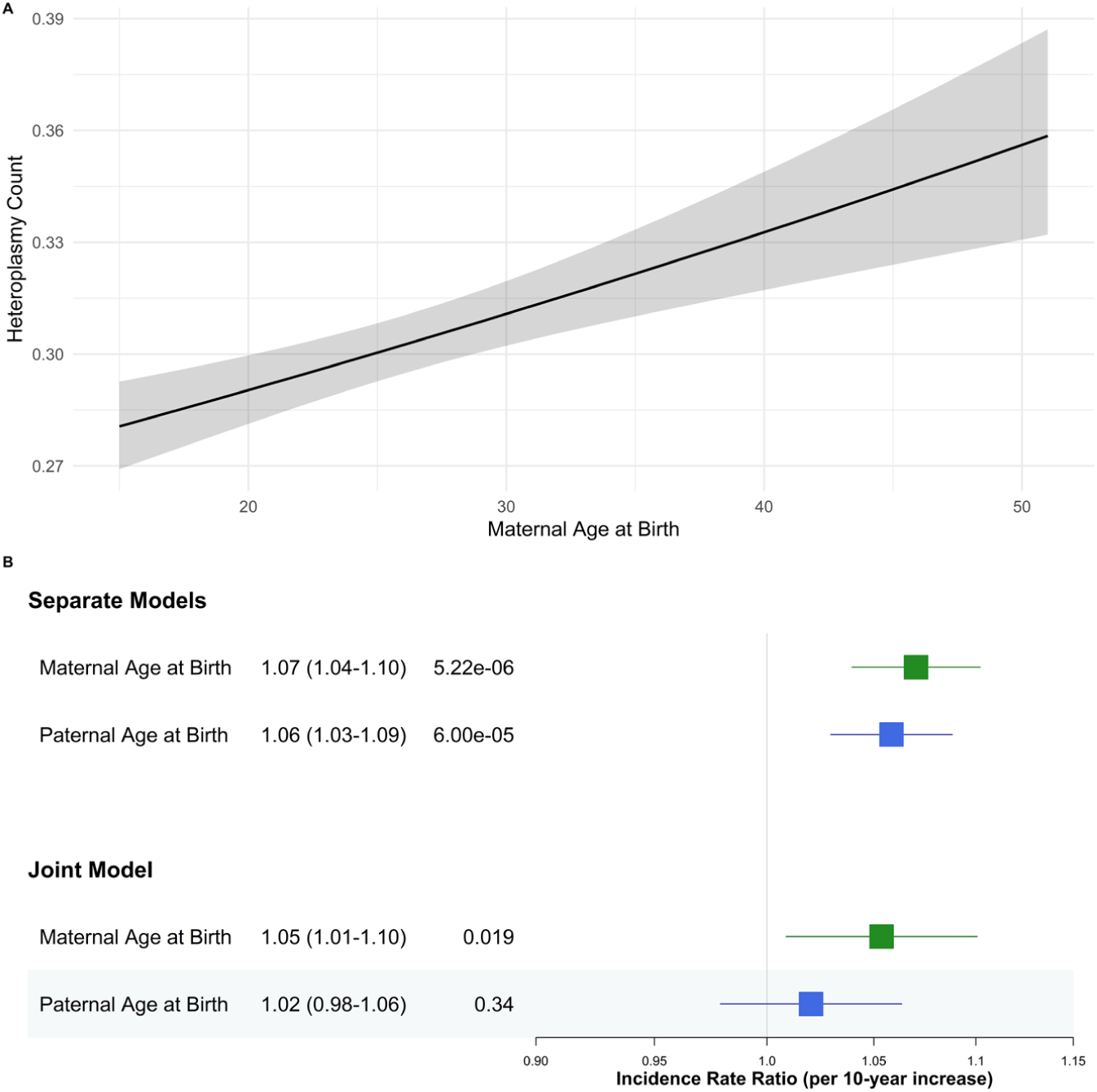
Impact of parental age on heteroplasmy count. **a,** Correlation between maternal age at birth and offspring heteroplasmy count. **b**, Forest plot showing the association of parental age in individual models (**top**) or in a joint model (**bottom**), with estimates scaled per 10 years. Analyses are adjusted for participant age, sex, and smoking exposure.

We also calculated shared and not shared modified MLC sum scores (mMSS), which capture the overall predicted deleterious impact of heteroplasmies within an individual.^8,17^ Prior work has demonstrated that higher mMSS is associated with overall mortality, as well as various aging-related phenotypes, including risk for hematological cancers.^5,7–9,18^ Surprisingly, despite maternal age at birth being associated with higher heteroplasmy count, we do not see an association with mMSS (P=0.28), suggesting that these additional heteroplasmies are largely benign.

### Overall mMSS is largely determined by not shared variants

Given the strong predictive role of mMSS in cancer and aging, we wanted to determine what proportion of the overall mMSS is explained by shared vs. not shared heteroplasmies, as this could have important implications for prevention – if most of the risk associated with mMSS is driven by not shared variants (largely de novo), then development of strategies to reduce or prevent heteroplasmy acquisition could have clinical utility. We observed that the vast majority of the signal for the overall mMSS is due to not shared mutations (Pearson’s r = 0.94, r^2^ = 0.88, **Figure 4**), consistent with most of the risk associated with mMSS driven by somatically acquired mutations.

**Fig. 4.**
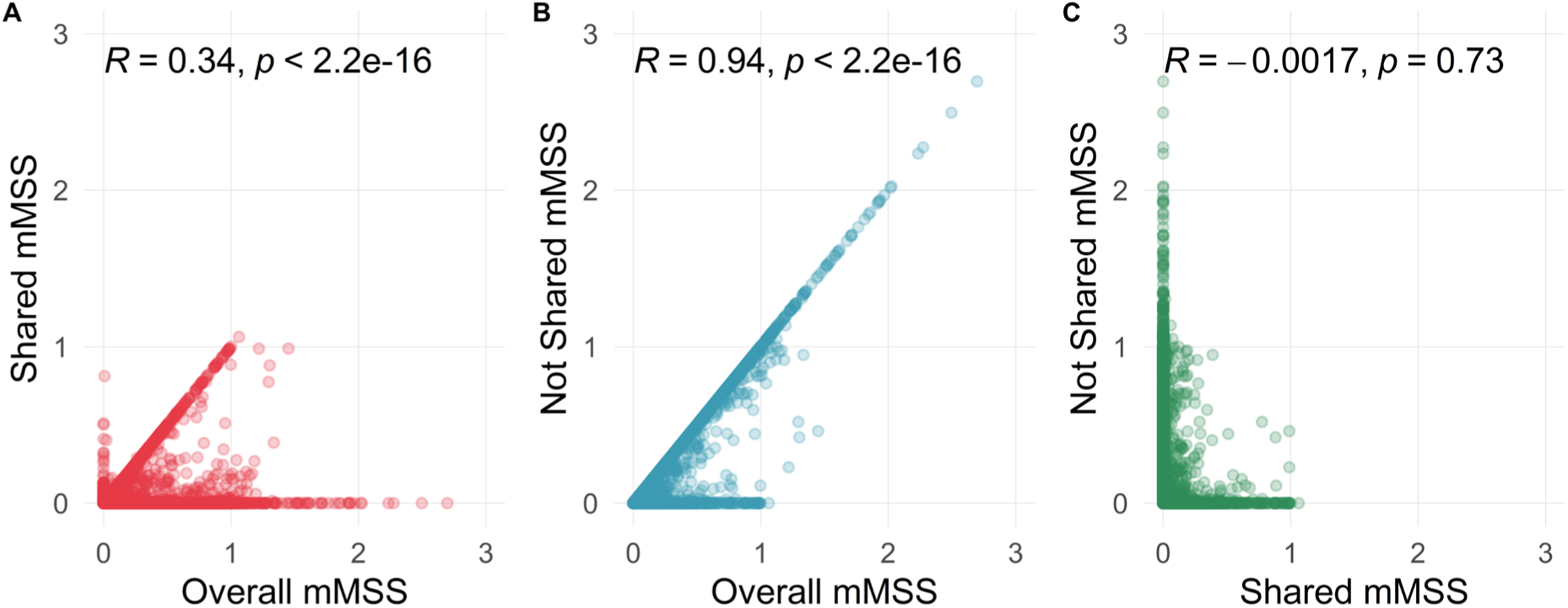
Correlation between overall mMSS and sharing status stratified mMSS. **a,** overall mMSS vs. shared mMSS; **b,** overall mMSS vs. not shared mMSS; **c,** shared mMSS vs. Not shared mMSS. Pearson’s correlations (R) are displayed.

### Distribution of mMSS by relative pair

The ability to identify shared heteroplasmies is a function of the relationship pair, with MZ Twins sharing both heteroplasmies inherited from Mother and germline de novo variants. In contrast, for Full Siblings, the mother has to transmit the same heteroplasmy to both children for it to be shared. Thus, we expect that shared mMSS, which is a function of the number of heteroplasmies weighted by their predicted functional impact, should differ by relative pair, which is indeed what we observe (**Table 2**).

**Table 2:** Shared mMSS Stratified by Relationship.

| mMSS Category | MZ Twins | Mother-Offspring | Full Siblings |
| --- | --- | --- | --- |
| 0 | 288 (84.7%) | 6970 (89.1%) | 33324 (92.2%) |
| >0-0.10 | 32 (9.4%) | 554 (7.1%) | 1860 (5.1%) |
| >0.10-0.25 | 12 (3.5%) | 196 (2.5%) | 669 (1.9%) |
| >0.25 | 8 (2.4%) | 100 (1.3%) | 295 (0.8%) |

Values shown as N (%).

As expected based on the individual heteroplasmic variant data (**Figure 2**), not shared mMSS has significantly higher values than shared mMSS (**Figure 5A**). We also observed an association between haplogroup and shared mMSS (overall P<0.003), largely driven by non-European haplogroups (A, P<2×10^−6^; L1 P<0.002, **Supplementary Table 2**). Thus, we limited downstream analyses to participants with European ancestry haplogroups and included principal components (PCs) generated from mitochondrial SNPs to correct for any potential residual population substructure (see **Methods**, **Supplementary Figure 3**).

**Fig. 5.**
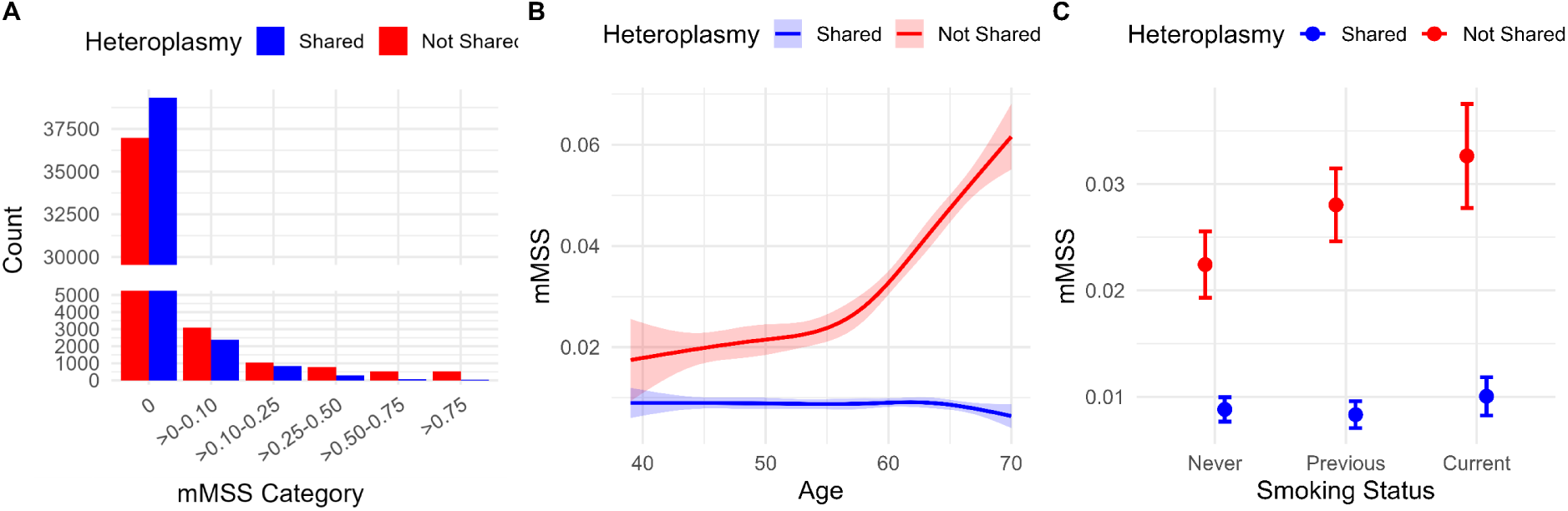
mMSS characteristics stratified by inheritance. **a,** categorical mMSS. **b,** mMSS as a function of age. **c,** mMSS as a function of smoking status. Age and smoking status were run in the same regression model and are additionally adjusted for sex, relative type, and top 4 mitochondrial PCs. Blue indicates shared mMSS and red indicates not shared mMSS.

Consistent with the shared mMSS being driven by inherited heteroplasmies (with the exception of germline de novo mutations in the MZ Twins, which are also shared), we observed no significant association with age (**Figure 5B**) or smoking status (**Figure 5C**), whereas not shared mMSS is significantly associated with both measures.

### Impact of shared vs not shared variants on overall mortality and hematological malignancies

A major question about the role of heteroplasmy in aging-related disease revolves around the question of causality. One way to address this issue is through Mendelian randomization using the shared mMSS as an instrument variable. Since the shared mMSS is present from conception and thus, it’s not merely a marker of clonal expansion, if the mMSS is associated with an outcome, that would provide strong evidence for a causal role. Thus, we assessed the association of shared and not shared mMSS with overall mortality, hematological malignancies, leukemia, and myeloid neoplasms (MN) using robust methods to account for the related pairs (see **Methods**). Overall, we saw highly concordant results between the shared mMSS and not shared mMSS, and the shared mMSS was significantly associated with hematological malignancies (**Figure 6, Supplementary Figure 4**), providing evidence for a causal role of predicted deleterious heteroplasmies in risk for hematological malignancies. While not statistically significant, the point estimate for overall mortality was larger than the not shared mMSS (relative risk [RR] 1.65 vs 1.21), further supporting a functional role for aging-related disease.

**Fig. 6.**
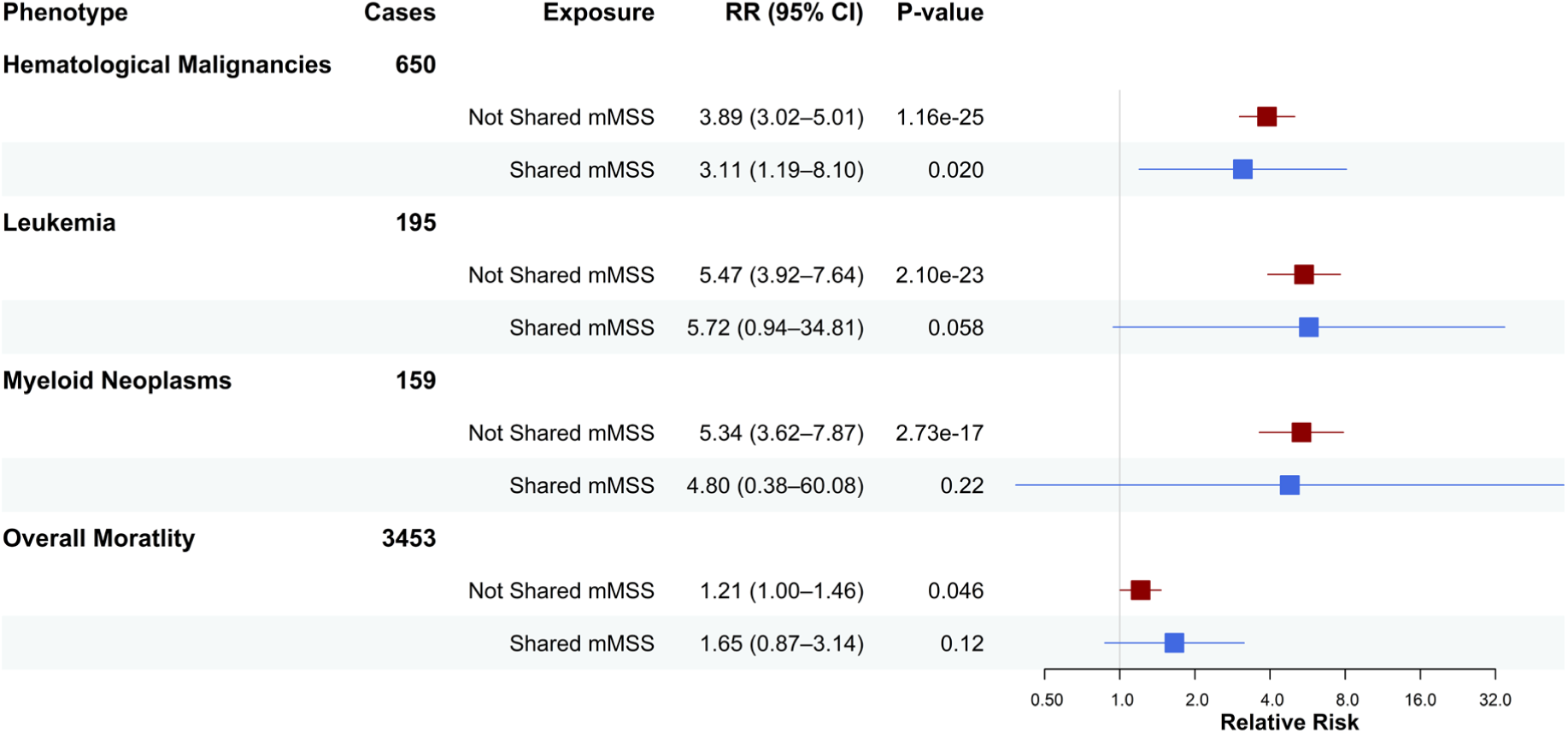
Forest plot depicting the association of shared and not shared mMSS with hematological malignancies and overall mortality. Analyses are adjusted for age, sex, relative type, smoking exposure, and the top 4 mitochondrial PCs. Red indicates de novo mMSS and blue indicates shared mMSS. RR = relative risk; CI = confidence interval. Hematological malignancies excludes multiple myelomas, which we have previously shown are not associated with heteroplasmy^5^

### Impact of shared vs not shared variants on spliceosome CHIP

We have previously observed a significant enrichment of higher mMSS levels in individuals with spliceosome CHIP mutations compared to other M-CHIP mutations.^8^ Spliceosome CHIP mutations are high-risk, and thus, one possible explanation could be that stronger CHIP driver mutations allow for more deleterious mitochondrial heteroplasmies to be clonally expanded as passenger mutations. Alternatively, deleterious heteroplasmies can promote spliceosome CHIP acquisition and/or clonal expansion. Consistent with the latter hypothesis, we observe a significant association between both the shared and not shared mMSS and the presence of high-risk M-CHIP and spliceosome mutations (**Figure 7**). This is in striking contrast to low-risk M-CHIP, in which we only see a significant association with not shared mMSS, despite a much larger number of low-risk CHIP carriers.

**Fig. 7.**
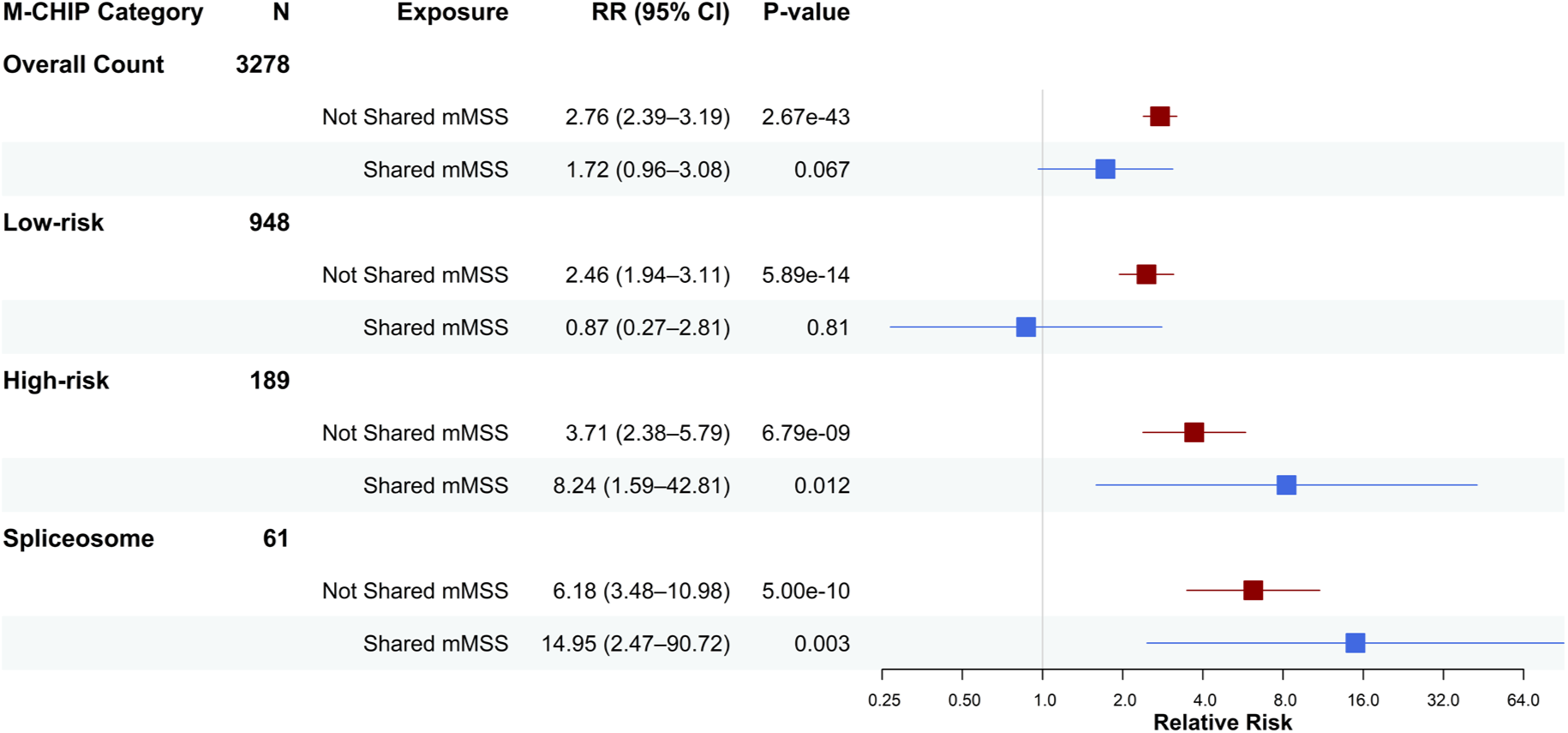
Forest plot depicting the association of shared and de novo mMSS with CHIP mutations. Analyses are adjusted for age, sex, relative type, smoking exposure, and the top 4 mitochondrial PCs. Low-risk = single *DNMT3A* mutation; High-risk = *JAK2*, *SF3B1*, *SRSF2*, *ZRSR2*, *U2AF1*, *FLT3*, *IDH1*, *IDH2*, *NPM1*, *RUNX1*, *TP53*; Spliceosome = *SF3B1*, *SRSF2*, *U2AF1*, *ZRSR2*. Red indicates de novo mMSS and blue indicates shared mMSS. OR = odds ratio; CI = confidence interval.

### Validation in the All of Us Research Program cohort

After generating heteroplasmy calls and data cleaning (**Methods**), we identified 13,831 first-degree relative pairs in the All of Us cohort (9,113 Mother-Offspring, 4,718 Full Sibling; note MZ Twins were excluded due to potential sample handling issues, see **Methods**) (**Supplementary Table 3**). We observed broadly similar results to the UKB data for shared vs. not shared heteroplasmies, with similar correlations of VAF for relative pairs (**Supplementary Figure 5a**), higher VAF for shared heteroplasmies (**Supplementary Figure 5b**), higher likelihood of mutation severity for not shared heteroplasmies (**Supplementary Figure 5c,d**) and mMSS (**Supplementary Figure 5e**), and a strong association of age with mMSS for not shared heteroplasmies, but not for shared heteroplasmies (**Supplementary Figure 5f**). Given the much more extensive genetic diversity of the All of Us cohort, we did not exclude individuals based on genetic ancestry, but included both continental haplogroup (EA, AF, AS) and mitochondrial PCs in the analyses to account for mitochondrial genetic ancestry. We assessed the association of shared and not shared mMSS with overall mortality and hematological malignancies, and observed qualitatively consistent effects with UKB results for the hematological categories, though effect sizes were noticeably smaller for both shared and not shared mMSS, with not shared mMSS effect estimates all significantly smaller than in the UKB (all P<0.01) (**Supplementary Figure 6a**). Excluding individuals >70 years of age, to match the UKB upper age cut-off, we observed larger effect estimates that were no longer significantly different from the UKB, and again found consistent effect sizes for shared and not shared mMSS (**Supplementary Figure 6b**). Combining both the UKB and the age-restricted All of Us results, we observe a significant association of shared mMSS with overall hematological malignancies (RR=2.81, 95% CI 1.29-6.13, P<0.009).

We also looked to validate the association of shared mMSS with high-risk CHIP. There was a substantially lower CHIP rate in the All of Us cohort compared to UKB (2.1% vs. 7.8%), likely reflecting a combination of lower average age in the All of Us cohort, in addition to reduced sensitivity to detect CHIP from whole-genome sequencing (WGS) data (**Supplementary Tables 1 and 3**). Among 24 high-risk CHIP carriers, we observed directionally consistent results (RR=3.42, 95% CI 0.28-41.6, P=0.34), with no association seen for low-risk CHIP (**Supplementary Figure 7**). Combining both the UKB and the All of Us results, we observe a significant association of shared mMSS with high-risk CHIP (OR=5.81, 95% CI 1.44-23.4, P<0.014).

### Identification of recurrent mitochondrial mutations associated with hematological malignancies

A hallmark of driver mutations in cancer is the association of inherited or recurrent mutations. We therefore assessed the effect of individual variants, including both homoplasmies and heteroplasmies, on hematological malignancy risk in self-reported white British ancestry UKB participants (see **Methods**). We identified 5 statistically significant ultra-rare variants (<1/10,000 carriers) after Bonferroni correction for the number of variants tested (**Figure 8**, **Supplementary Figure 8a, Supplementary Table 4**). These variants were only observed as low VAF heteroplasmies (**Supplementary Table 5**), suggesting that they are de novo events. We attempted to validate results in the non-white British UKB participants and in the All of Us Research cohort. Two single base pair deletions had consistent effect directions and were significant in the non-white British UKB participants (11031GA>G, 12417CA>C), with 11031GA>G also significant in the All of Us Research cohort (**Figure 8, Supplementary Figure 8b,c**). 11031GA>G occurs in the *ND4* gene in a region with high mMLC scores for missense mutations (mMLC >0.65 for missense mutations at position 11031 and flanking bases), suggesting that mutations in this region are likely to be highly deleterious. 12417CA>C is similarly found in a region intolerant to missense mutations (mMLC >0.35 for missense mutations at position 12417 and flanking bases) in the *ND5* gene. Both deletions also disrupt the open reading frames leading to premature stop codons, providing strong evidence for deleterious impacts on mitochondrial function.

**Fig. 8.**
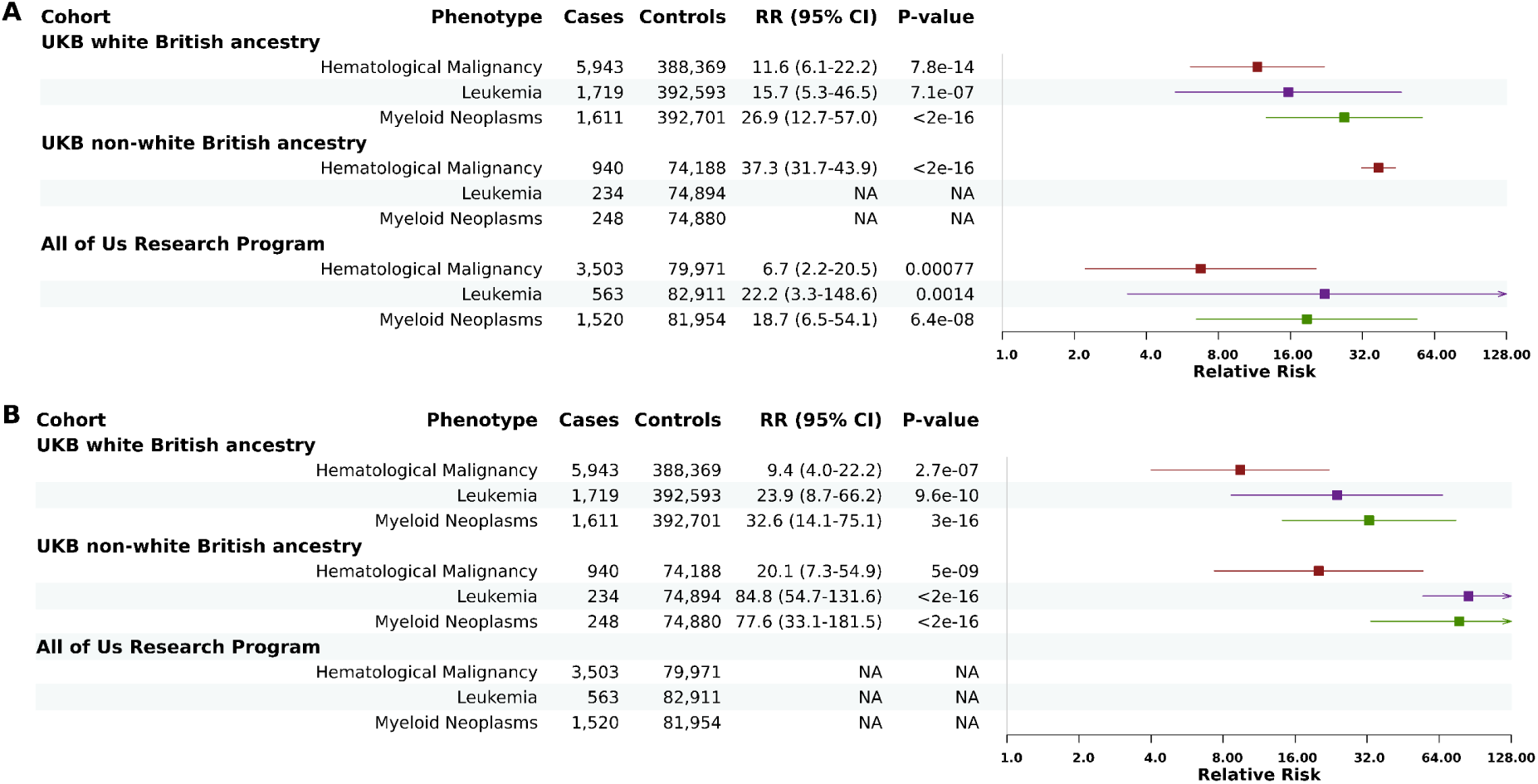
Forest plot depicting validated individual mtDNA variant associations with hematological malignancies. **a,** 11031GA>G**, b,** 12417CA>C. Analyses were adjusted for age, sex, smoking exposure, mitochondrial continental haplogroup, and mtDNA PCs. “NA” denotes that no cases carried the variant.

## Discussion

The role of mitochondrial heteroplasmy in aging-related disease, and in cancer in particular, has been increasingly recognized. The mitochondrion is a highly-evolved organelle for energy production and distribution, but it is also essential in a broad array of cellular functions critical in cancer development, including apoptosis and innate immunity signaling.^19,20^ These additional mitochondrial functions are key players in tumorigenesis, providing for cellular adaptation to the environment resulting in flexibility for tumor cell growth and survival in otherwise harsh environments (e.g. nutrient depletion, hypoxia, and cancer treatments).^21^ Evidence has begun to emerge for a causal role, including a recent study from Boscenco and colleagues that identified hotspot mutations in mitochondrial rRNA genes associated with cancer risk.^10^ Recurrent mutations is a hallmark of causal variants, and the identified hotspot mutations were further shown to have a dominant negative effect with heteroplasmy as low as 10% VAF. In the context of hematological cancers, while predicted deleterious heteroplasmies are associated with risk for MN and CLL,^8,9^ no hotspot mutations were previously identified, leaving open the question of whether mtDNA heteroplasmy is causal or largely driven by passenger mutations in the context of CHIP and other unobserved nuclear DNA clonal hematopoiesis drivers, as suggested by a recent GWAS of heteroplasmic mtSNV burden.^6^ The question of causality has significant implications for prognostication and therapeutic strategies.

Leveraging the UKB, a large population-based cohort, we were able to identify ∼22,000 first-degree relative pairs, allowing us to dissect mitochondrial heteroplasmy into shared (inherited) and not shared (largely de novo) components. We were able to confirm a bottleneck size of ∼3 mtDNA units, consistent with prior findings^16^, and demonstrate high VAF concordance across relative pairs, consistent with the bottleneck size and the number of bottlenecks separating the relative pairs. Likewise, we were able to confirm prior observations of increased overall heteroplasmy with increasing maternal age^22^, which has also been explicitly linked to increased de novo heteroplasmy.^23^ Surprisingly, we did not see a significant increase in predicted deleterious heteroplasmy burden (mMSS), suggesting that despite some relaxed constraint on de novo heteroplasmy with maternal age, natural selection is still acting to suppress more deleterious heteroplasmies.

As expected, shared and not shared heteroplasmies had strikingly different VAF profiles, and shared heteroplasmies demonstrated clear evidence for selection, with reduced levels of functional and/or predicted deleterious variants relative to not shared variants. By separating shared and not shared heteroplasmies, we were able to demonstrate that the mMSS, which is associated with a significant increase for MN/CLL risk (hazard ratio ∼4-5 per unit change of mMSS),^5,8,9^ is largely driven by not shared variants. This has important implications from a prevention standpoint, as lifestyle changes can modify risk of heteroplasmy acquisition (e.g., reduction of smoking exposure), and further work is warranted to see if specific drugs may reduce or prevent heteroplasmy acquisition and/or expansion in those already at high risk for MN/CLL development.

To address the question of whether deleterious mitochondrial heteroplasmies are causal for hematological cancer, we relied on the shared mMSS, which is fixed at conception and thus temporally upstream of CHIP and other potential confounders, as an instrument variable for Mendelian randomization. We found remarkably consistent effect estimates between the shared and not shared mMSS, and a statistically significant association for the shared mMSS and risk of hematological cancer, with consistent effects in both the UKB and All of Us cohorts. These results provide strong evidence for a causal role for predicted deleterious heteroplasmies and hematological cancer risk. Combined with the observation that the vast majority of the mMSS (∼90%) is somatically acquired, and that mMSS has a causal role in hematological cancer, this opens up new areas for both prevention and treatment.

We were also able to show that shared mMSS was associated with high-risk CHIP, and in particular, spliceosome CHIP. This is consistent with our prior study showing a significant enrichment of mMSS in spliceosome M-CHIP relative to other M-CHIP, and may, in part, explain the synergistic effect on MN risk that was observed between mMSS and M-CHIP.^8^ This result argues that while heteroplasmy may also be a consequence of CH due to nuclear variants, predicted deleterious mitochondrial heteroplasmies directly contribute to development of specific forms of CHIP. Since CHIP is a function of both nuclear DNA mutation (acquisition) and selection (clonal expansion), mitochondrial heteroplasmy must directly impact one or both of these mechanisms. Given the specificity for spliceosome CHIP, it seems more plausible that predicted deleterious heteroplasmy is contributing to clonal expansion (no known mechanism would explain mitochondrial dysfunction contributing to gene-specific mutations in the nuclear genome). This potential explanatory mechanism is supported by prior work in longitudinal samples demonstrating that heteroplasmies with higher mMSS are more likely to clonally expand.^18^

Finally, we assessed whether individual mtDNA variants are associated with hematological malignancies, a signature of cancer driver mutations. We identified 2 ultra-rare single base pair deletions, which displayed the hallmarks of recurrent de novo mutations (no homoplasmic carriers, low VAF, highly conserved genic region, disruption of open reading frame), one of which was significantly associated in all 3 independent cohorts (white British UKB, non-white British UKB, and All of Us Research Program). These deletions had remarkably large effect sizes, similar to high-risk CHIP mutations, providing strong support for a causal role in hematological malignancies. Notably, the recurrent deletions were located within the *MT-ND4* and *MT-ND5* genes, which encode core membrane subunits of mitochondrial respiratory chain complex I. Rare variants in mitochondrially encoded complex I components have been found in cancer, including acute myeloid leukemia.^24,25^ The fact that both variants are predicted truncating mutations affecting highly conserved coding regions further supports a functional role, suggesting that partial complex I dysfunction may represent a previously unrecognized mechanism promoting clonal expansion and malignant transformation in hematopoietic cells. Moreover, the presence of complex I variants may results in non-random selection of hematopoietic clones with somatic mutations in nuclear DNA shaped by metabolic selective pressures.^26^ This could explain the significant association between deleterious heteroplasmies and spliceosome mutations.

The current manuscript has several limitations. First, Full Siblings make up the vast majority of the relative pairs, and given that for a heteroplasmy to be shared, it would have to be transmitted from the mother to both siblings. Thus, a subset of heteroplasmies characterized as not shared in these pairs are in fact inherited but not shared. Thus, we likely underestimate shared mMSS in these individuals. Given that not shared mMSS is strongly associated with our phenotype of interest, this likely contributes to false-negative results rather than false-positive results. Second, due to selection, the shared mMSS, which is largely inherited, has a narrower range than the not shared mMSS, with many more individuals with low or zero mMSS. Thus, despite a large sample size, power is somewhat limited to test the associations of shared mMSS with hematological cancer subtypes. Despite the reduced power, while not significant, the remarkably similar effect sizes for shared and not shared mMSS across the hematological cancer subtypes and overall mortality provide confidence in the overall finding of causality. Third, the UKB, which drives most of our results, has a healthy volunteer bias and is limited to participants with European haplogroups. While the All of Us Research Program, which is a much more genetically diverse population, demonstrated directionally consistent effects, additional studies are warranted to comprehensively extend findings to non-European ancestry populations. Together, these analyses implicate predicted deleterious mtDNA heteroplasmies as causal contributors to hematological cancer risk, while underscoring that most of the risk is somatically acquired. This framework lays the groundwork for efforts to identify modifiable exposures and interventions that limit heteroplasmy acquisition or selectively counteract its functional consequences in high-risk individuals.

## Methods

The current study was approved by the Johns Hopkins Medicine Institutional Review Board.

### Study populations

*UK Biobank.* The UK Biobank (UKB) is a population-based prospective study of ∼500,000 participants between the ages of 40 to 69 years recruited from across the United Kingdom from 2006 to 2010 ^27^. Extensive information is available on demographics and lifestyle factors, along with whole-genome sequencing (WGS) data, which is used to assess mitochondrial variation (see below). Sex is based on self-report (FIELD ID 31), and age is the participant age at study entry (FIELD ID 21022). The data is linked to the death registry, cancer registry, hospital admissions, and primary care visit data, which are used to assess overall mortality and cancer status.

*All of Us.* The All of Us Research Program is an on-going population-based prospective cohort study of adults (18 and older) recruited across the US since 2017.^28^ The study focuses on recruiting diverse populations for race, ethnic background, age, sex, gender identity, and sexual orientation. Participants provided consent to respond to survey questions, link electronic health records, and to provide biospecimen for genetic analysis, including WGS. Cancer outcomes are identified through electronic health records. Death records are identified from the “aou_death” table, which is derived from electronic health records and HealthPro (see https://support.researchallofus.org/hc/en-us/articles/10237876482196-How-to-find-participant-death-reports).

### Identification of related individuals

The KING software package (version 2.3.2) was run using autosomal SNPs obtained from the genotyping array (UKB) and WGS (All of Us) data with MAF ≥0.02 (**Supplementary Figure 9**). After running KING, we randomized the which sample ID was ID1 vs ID2, as the original genotype file is non-random with respect to disease status. For participants related to multiple individuals, we only retained a single pair of related individuals using the following priority for retention: MZ > Mother-Offspring > Full Siblings. Note that Father-Offspring pairs were only used as negative controls for inherited (shared) mitochondrial variants. We also removed any Parent-Offspring pairs with an age difference of <15 years (UKB n=6, All of Us cohort n=161) and any Full Sibling pairs with age difference >40 (n=2 for All of Us cohort).

In the All of Us cohort, we identified 2,640 duplicate/MZ twin pairs, which was well in excess of what was expected given the cohort size. Upon preliminary quality control, we observed multiple anomalous clusters in which individual samples were flagged as MZ twins with every other sample in the cluster, despite having distinct dates of birth and unique visit dates. The largest such cluster comprised 25 samples. We suspect these are either sample contamination or technical duplicates rather than true biological relatedness. After consulting the All of Us Research Program support team and confirming that no method is currently available to distinguish genuine MZ twin pairs from sample handling error or duplicated samples, all pairs classified as duplicate/MZ by KING were excluded from subsequent analyses.

### mtDNA variant determination

*UKB*. mtDNA variation was determined from WGS generated from buffy coat DNA in 490,270 UKB participants using MitoHPC implementing the GATK Mutect2 variant detection setting,^29,30^ (version 20230418; all default settings with random down-sampling to use a maximum of 222K reads) as previously described.^4,5,9^ Briefly, we excluded samples (n = 3,776) if they met any of the following criteria: 1) Haplocheck^31^ contamination level ≥3%, suggestive of sample cross-contamination; 2) ≥2 variants mapping to the same known NUMT sequence; 3) ≥2 variants belonging to different mitochondrial haplogroups; or 4) low minimum base coverage (<100x) or low mean base coverage (<500x). We also removed 12,001 participants with low mitochondrial DNA copy number (mtDNA-CN ≤40), which can lead to false-positive heteroplasmy calls.^4^ Finally, we removed 699 participants with a heteroplasmy count >5, as these samples are not distinguishable from contaminated samples ^5^. Overall, 13,918 participants were excluded (some samples failed multiple checks), resulting in 476,352 participants with heteroplasmy information for downstream analyses.

*All of Us Research Program.* mtDNA variation was determined from WGS generated from whole blood and saliva-derived DNA using the same pipeline as described for UKB, using MitoHPC version 20240306. To assess whether DNA source (whole blood vs. saliva) might impact our analyses, we tested the association of mMSS with hematological malignancies in a random cohort, where we randomly selected 20% of all samples with WGS and EHR data (n=61,122; whole blood n = 56,583, saliva n = 4.539) drawn from the full All of Us cohort WGS dataset. We observed no significant difference in associations (**Supplementary Figure 10**), and thus all analyses included both sample source types.

We used a threshold of 5-95% variant allele fraction (VAF) to define heteroplasmic SNVs, with variants outside these thresholds considered homoplasmic.

### QC for heteroplasmic variants

Analyses, with the exception of the individual variant associations with hematological malignancies (see below), were restricted to SNVs, as insertions / deletions have higher false-positive calls.^4^ For SNVs, we excluded variants with read depth <300 and those flagged as base quality, mapping quality, strand bias, slippage, weak evidence, position, clustered, fragment length, and haplotype flags in the FILTER column of the VCF. We additionally excluded heteroplasmic variants at poly-C homopolymer regions (chrM 296-318,493-502,511-524,538-545,561-574,954-965,5888-5895,8269-8288,16178-16193). We also excluded mt12684G>A and mt12705C>T, as these were identified as false-positives due to their association with mtDNA copy number.^8,18^ In addition, we excluded 16093T>C, as this was identified as a false positive inherited (shared) variant in UKB Father-Offspring pairs 5 times, accounting for more than 50% of the false positives (5/9), and was the only variant observed more than once.

### Determination of shared vs. not shared heteroplasmy

For each heteroplasmy detected at VAF 5-95%, we assessed whether the variant was present in the related individual with a threshold of ≥3 alternate allele reads (i.e., no longer using the VAF 5-95% threshold to avoid edge effects). If detected at the new relaxed threshold, we deemed this a “shared” heteroplasmy, and if not, the heteroplasmy was deemed “not shared”.

### mtDNA PCA calculation

Mitochondrial DNA principal components (mtDNA PCs) were computed using all quality-controlled heteroplasmic and homoplasmic variants called by MitoHPC. Variants were converted to PLINK2 pgen format and filtered to retain only those with a minor allele frequeny greater than 0.01. The resulting filtered variant set was then used to compute mtDNA PCs via the *prcomp* function in R.

### Mitochondrial haplogroup definition

Continental mitochondrial haplogroups were defined based on the Simple mtDNA Tree from Mitomap (see Figure 7 from https://mitomap.org/foswiki/bin/view/MITOMAP/MitomapFigures).

### Mitochondrial variant annotation

All annotations are available as part of the MitoHPC pipeline (https://github.com/ArkingLab/MitoHPC). Note that the mitochondrial local constraint (MLC) score has been modified from the original publication^17^ to take into account homoplasmy count. As previously described, the modified MLC (mMLC) score is calculated as mMLC = MLC / 1 + (log_10_(homoplasmy_count +1)).^8^ The full annotation table can also be found under (https://github.com/ArkingLab/MitoHPC_Process/mito_genome_annotation.txt)

### CHIP Calling

WES/WGS CRAM files were aligned to hg38. Variant calling was performed using Genome Analysis Toolkit (GATK) v.4.2.2. Mutect2.^29,32^ Mutect2 was run in ‘tumor-only’ mode using non-default parameters: gatk Mutect2 --panel-of-normals ${ref_pon} --germline-resource ${ref_germ}. Raw variants called by Mutect2 were filtered out with FilterMutectCalls using the estimated prior probability of a reading orientation artifact generated by LearnReadOrientationModel.

Because of a known duplication in hg38, *U2AF1* is not reliably called by variant callers. Thus, we employed a pileup-based approach for *U2AF1* calling (https://github.com/weinstockj/pileup_region).^8,33^

### CHIP variant filtering

We only assessed genes that were part of a custom CHIP panel (**Supplementary Table 6**). Variant annotation was performed using ANNOVAR.^34^ Variant filtering was performed similarly to our previously described strategy.^8^ Briefly, we considered how likely a variant was to be pathogenic, an artifact, or germline. For variant pathogenicity, we used a previously published whitelist.^33^ To avoid artifactual variants, we applied, the Mutect2 FILTER of PASS, weak_evidence and germline, depth ≥ 20, alt count ≥3 (SNVs [single nucleotide variant]) or ≥5 (MNVs [multi nucleotide variant]), alt F1R2 ≥1, alt F2R1 ≥1 and VAF ≥2% (SNVs) or ≥10% (MNVs). In the case of variants present on the X chromosome, we divided the VAF by 2 in males. To avoid germline variants, we only included variants present in non-cancer populations (non_cancer_AF_popmax) under 0.001 in the gnomAD exome collection v2.1.1.^35^ Further, variants present at germline levels in ≥80% of the individuals they occurred in were removed. This latter filter was applied only in variants occurring in 3 or more individuals.

On a sample level, we excluded individuals with more than 3 mutations, all of which are represented by MNVs or individuals with more than 9 CHIP variants.

### Calculation of mtDNA bottleneck size

The size of the intergenerational bottleneck (*N_e_*) was estimated as previously described.^16^ Briefly, the method assumes that heteroplasmies follow the Kimura distribution^36,37^, and thus for 1 generation, *N_e_* = -1 / log(*b*), where b = 1 – var(*h*) / (*p* x (1-*p*)), where *h* is the mutation frequency in Offspring and *p* is mutation frequency in Mothers. To get sufficient mutations to estimate var(*h*), we binned heteroplasmies based on their VAF in the Mothers using intervals of 5%, ranging from 10-90% (16 bins total), and calculated *h* across all heteroplasmies in the Offspring in the interval and *p* was calculated as the mean VAF in Mothers in the interval. Following Arnadottir and colleagues,^16^ we excluded VAF<10% and >90% to ensure stable estimates. An overall weighted average *N_e_* was calculated based on the number of relative pairs in each interval. The 95% confidence was calculated as:

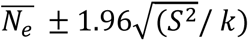, where 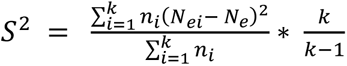, where *k* is the number of intervals (16), *n_i_* and *N_ei_* are number of relative pairs and the *Ne* estimate for interval *i*, respectively.

### Phenotype definitions

*UKB*. Hematological cancer and its subtypes, leukemia, and myeloid neoplasms, were defined using ICD codes from the cancer registry linked to the UK Biobank as detailed in **Supplementary Table 7**.^8^ Prevalent and incident disease were classified relative to the date of blood collection for DNA. Incident cases were censored as of 12/20/2022 or at the date of death, whichever came first. Mortality was defined using the UKB Death Registry and censored as of 12/22/2022.

Smoking exposure included two variables, Smoking Frequency and Smoking Duration. Smoking Frequency was generated by combining Past Smoking Frequency (FIELD ID 1249) and Current Smoking Frequency (FIELD ID 1239) as outlined in **Supplementary Table 8**. Smoking Duration was calculated separately for former and current smoking. For former smokers Smoking Duration was calculated as the difference between the age at which the participant stopped smoking (FIELD ID 2897) and the age at which they started (FIELD ID 2867). For current smokers this was calculated difference between the age at study entry (FIELD ID 21022) and the age at which they started smoking (FIELD ID 3436). To fill in missing Smoking Duration data, we performed imputation within three joint smoking frequency classes (FIELD ID 1249/ FIELD ID 1239) with >50 missing values: 1) Missing / “Yes, on most or all days”; 2) “Smoked on most or all days” / “No”; 3) “Smoked on most or all days” / “Only occasionally”. Imputation was performed using the *mice* function from the ‘*mice’* R package with the following parameters: method = “midastouch”, m = 1, maxit = 1, seed = 1.

*All of Us Research Program*. Hematological cancer and its subtypes, leukemia, and myeloid neoplasms, were defined using SNOMED and ICD codes as detailed in **Supplementary Table 9**. Sex is based on self-identified “sex_at_birth” (including only “Female” or “Male” participants), sample source (whole blood vs. saliva) is from the “sample_source” column, and center is from the “site_id” column. Age is calculated using date of birth and visit date for each sample. Genetic ancestry is based on the “ancestry_pred_other” column in the ancestry_pred auxiliary file. For smoking we used concept id 1585857 (“Have you smoked at least 100 cigarettes in your entire life? (There are 20 cigarettes in a pack.”)

### Statistical analysis

To assess the association of mutation consequence and mMLC with shared vs not shared status, we used logistic regression with heteroplasmy type as a binary outcome, with mMLC treated as a continuous measure. For the relative pair type association with heteroplasmy count, we used negative binomial regression models to model the count data (glm.nb from the ‘*MASS’* R package), and the significance of relative pair type was assessed using ANOVA of nested models with and without relative pair type and adjusting for sex, age with a natural spline with 4 degrees of freedom (d.f.) to allow for non-linear associations, and smoking exposure.

For analyses of maternal age at birth and heteroplasmy, we used negative binomial models for heteroplasmy count and linear models for mMSS, adjusting for age (with natural spline with 4 d.f.), sex, and smoking exposure. Parental age at birth was calculated as the difference of parental age (FIELD ID 1845 / 2946) from participant age at study entry. Exclusions included if the age difference between parent and offspring was <15 years or if either maternal or paternal age at birth were missing. Note that parental age is only available for parents alive at time of study entry.

To test the association of haplogroup with mMSS, we used an ANOVA to compare multivariable linear regression model with and without haplogroup in the model, adjusting for age (natural spline with 4 d.f.), sex, relative pair type, and smoking exposure. Associations with age and Smoking Status (“Never”, “Previous”, “Current”) were limited to individual with European Ancestry haplogroups, and the regression models included age (natural spline with 4 d.f.), sex, relative pair type, and the top 4 mitochondrial PCs.

Associations with overall mortality were evaluated using multivariable Cox proportional hazards models, with time from DNA collection to death or end of follow-up (administrative censoring on December 20, 2022), whichever occurred first. Models were adjusted for age (natural spline with 4 d.f.), sex, Smoking Frequency, Smoking Duration, relative pair type, and top 4 mitochondrial PCs. In addition, participants were clustered by relative pair (‘cluster’ function) to account for relatedness. For overall hematological malignancies, incident cases were analyzed using Cox proportional hazards models as above. Prevalent cases were analyzed separately using log-binomial regression, with robust SEs calculated clustering on relative pair using the ‘*Sandwich’* R package vcovCL function. Incident and prevalent results were combined using inverse-variance fixed effect meta-analysis as implemented in the R ‘*metafor’* package. For leukemia and MN, prevalent and incident events were jointly analyzed due to the smaller number events using a log-binomial regression, with robust SEs calculated as above.

For analyses of overall M-CHIP count, we used a negative binomial model, and for low-risk, high-risk, and spliceosome M-CHIP we used log-binomial regression models testing for the presence/absence of a mutation. All analyses were adjusted for age (natural spline with 4 d.f.), sex, smoking exposure, relative pair type, and top 4 mitochondrial PCs.

Meta-analyses between UKB and All of Us cohort results used random effect models as implemented in ‘*metafor’* package using method = “REML”.

Individual mtDNA variant associations with hematological cancers were tested using logistic regression models implemented in REGENIE v4.1 with approximate Firth likelihood ratio test p-values. Analyses were adjusted for age and age^2^, sex, UKB intake center, imputed smoking duration, the top 10 nuclear PCs, the top 10 mitochondrial PCs, and mitochondrial continental haplogroup. Primary analyses were run in the white British ancestry individuals (“in.white.british.ancestry” = 1) with genotype encoded as a continuous variable between 0 and 1 to include heteroplasmic VAF. Indels were included in these analyses. No filters were applied to homoplasmic variants called by MitoHPC. Heteroplasmic variants were filtered out using the same filters used in our previous analyses. Indels had the same filters applied except for the slippage flag, which may be too stringent for low VAF heteroplasmic indels that are otherwise supported by sufficient alternate read depth. All variants were filtered to have a minimum VAF of 0.05 in a sample. A minimum allele count (MAC) filter of 10 across the entire UKB cohort was applied to variants that were run. Significant variants were determined after applying Bonferroni correction for the number of variants tested within a given phenotype (P < 0.05 / 9127 = 5.5×10^−6^). Selected variants were validated using log-binomial regression with genotype data recoded as 0/1, with heteroplasmies coded as 1, to obtain relative risk estimates and to validate in the UKB non-white British cohort and the All of Us Research Program. These models tested the variants jointly along with sex, age (natural spline with 4 d.f.), imputed smoking duration, the top 10 mitochondrial PCs, and mitochondrial continental haplogroup. Samples in the All of Us cohort were filtered to age ≤70 years. Covariates used in the All of Us cohort were age (natural spline with 4 d.f.), sex, smoking status, the top 10 mitochondrial PCs, and mitochondrial continental haplogroup.

## Data Availability

UK Biobank data is available through application to the UK Biobank (Application Number 17731). Access to the UK Biobank data can be requested at https://www.ukbiobank.ac.uk/enable-your-research/apply-for-access.

Access to All of Us Research Program data can be requested at www.researchallofus.org/.

## Code Availability

Code for data cleaning and analysis is available on our github repository: https://github.com/ArkingLab/mito_related.

Documentation on MitoHPC pipeline for DNA Nexus server is available in https://github.com/ArkingLab/MitoHPC/blob/main/docs/DNAnexus_CLOUD.md.

Documentation on extracting Mitochondrial and NUMT reads from Google Cloud is available in https://github.com/ArkingLab/MitoHPC/blob/main/docs/GOOGLE_CLOUD.md. Source data are provided with this paper.

MitoHPC heteroplasmy data processing scripts: https://github.com/ArkingLab/MitoHPC_Process

CHIP calling and cleaning pipeline: https://github.com/ArkingLab/CHIP_pipeline

## Acknowledgements

This research was conducted using the UK Biobank Resource under Application Number 17731. This work was supported by National Institutes of Health (NIH) grants R01AG085753 (D.E.A.), R01HL156144 (L.P.G.) and Department of Defense HT9425-25-1-0670 (L.P.G.). The content is solely the responsibility of the authors and does not necessarily represent the official views of the NIH. This manuscript is the result of funding in whole or in part by the National Institutes of Health (NIH). It is subject to the NIH Public Access Policy. Through acceptance of this federal funding, NIH has been given the right to make this manuscript publicly available in PubMed Central upon the Official Date of Publication, as defined by NIH.

---

**Supplementary Fig. 1.**
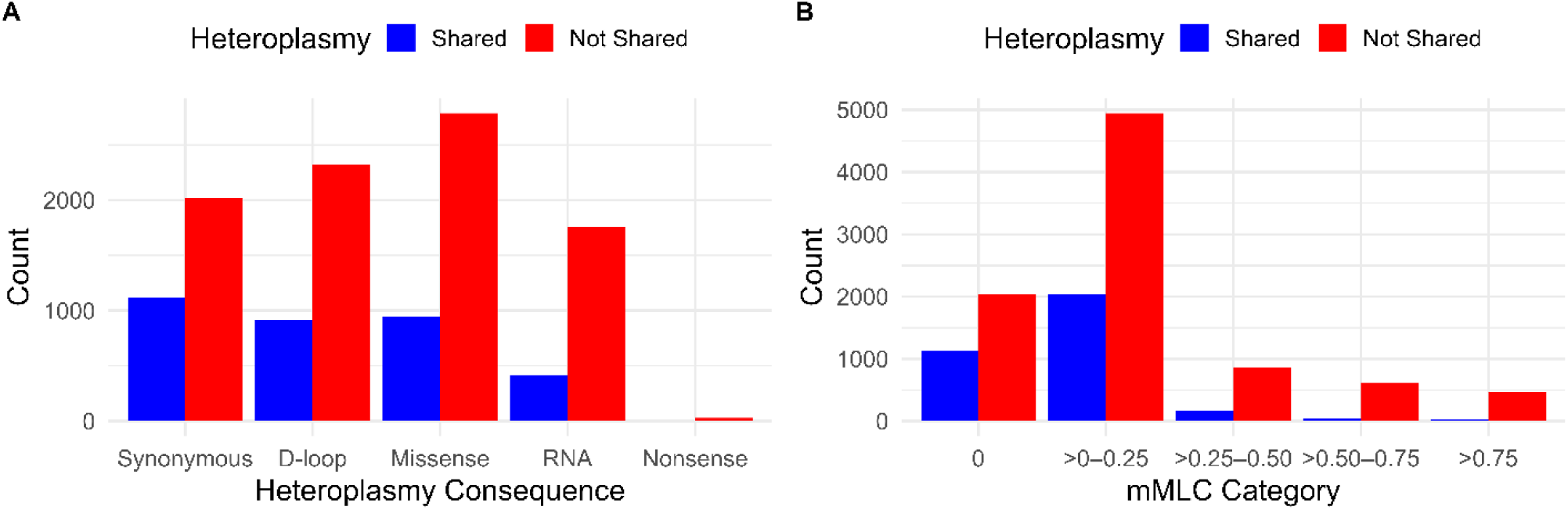
mtDNA heteroplasmy characteristics stratified by inheritance. **a,** Count of heteroplasmies stratified by mutational consequence (synonymous, missense, nonsense) or for non-coding variation, complex (D-loop vs RNA). **b,** Count of heteroplasmies stratified by mMLC categories. Blue indicates shared heteroplasmies, and red indicates not shared heteroplasmies.

**Supplementary Fig. 2.**
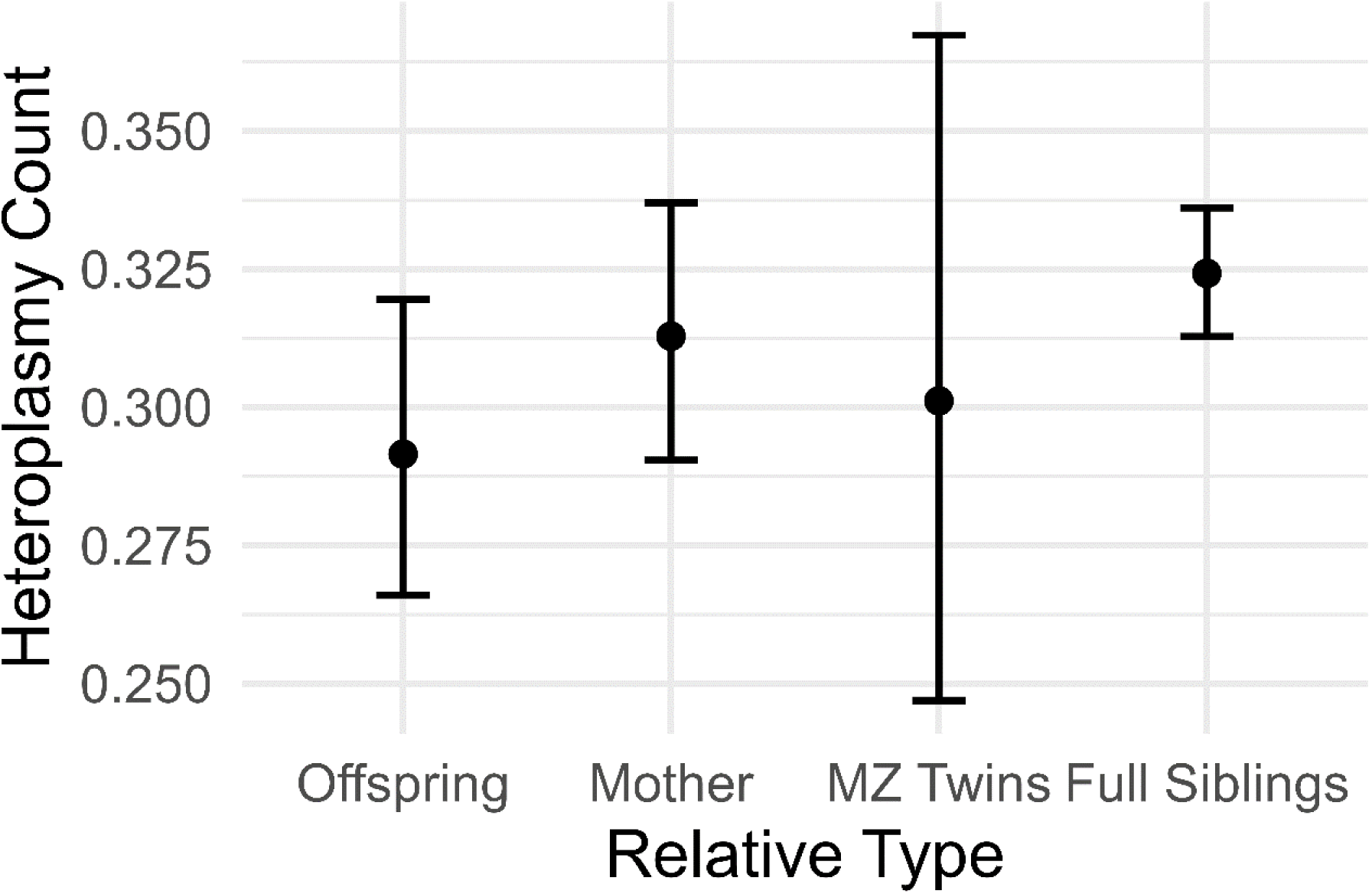
Heteroplasmy count stratified by relative type. The results are adjusted for age, sex, and smoking exposure.

**Supplementary Fig. 3.**
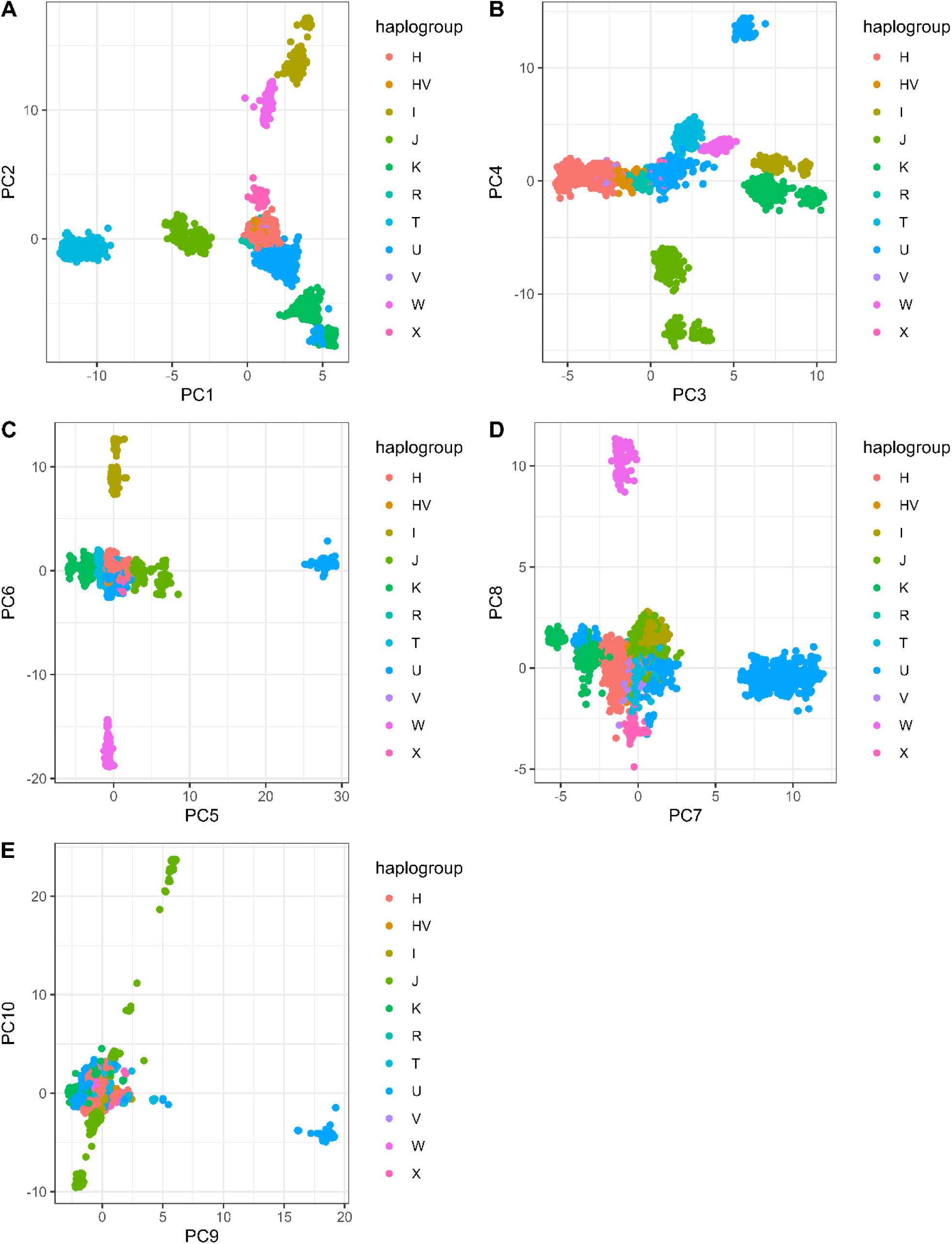
Plot of mitochondrial derived prinicpal components (PC) colored by haplogroup. PCs are ordered by variance explained, with PC1 explaining the most variance. **a**, PC1 vs PC2. **b**, PC3 vs PC4. **c**, PC5 vs PC6. **d**, PC7 vs PC8. **e**, PC9 vs PC10.

**Supplementary Fig. 4.**
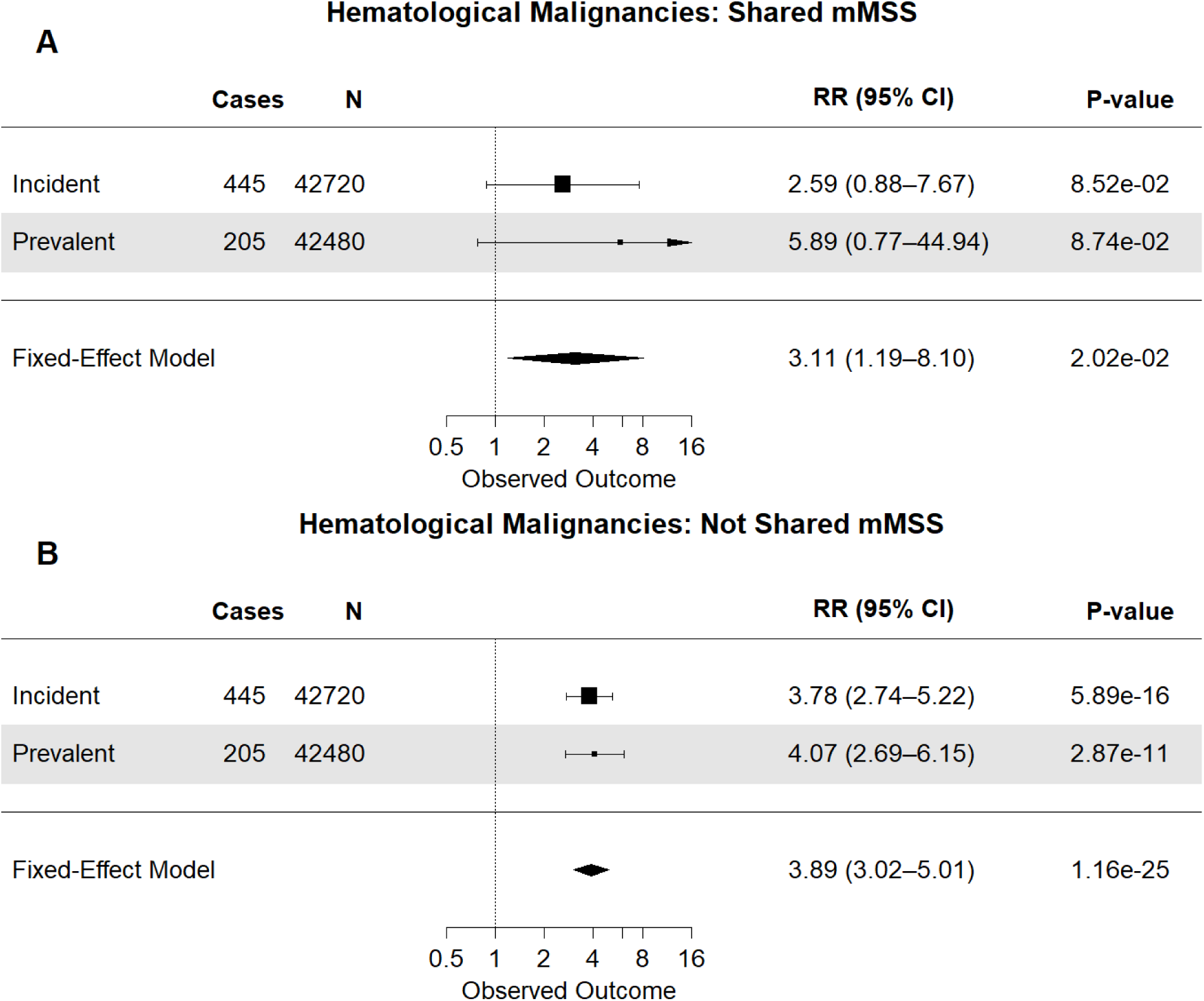
Forest plot depicting the association of a) shared and b) not shared mMSS with prevalent and incident hematological malignancies. Analyses are adjusted for age, sex, relative type, smoking exposure, and the top 4 mitochondrial PCs. RR = relative risk; CI = confidence interval. Leukemia and MN are not depicted, as the analyses for those phenotypes was conducted for prevalent and incident phenotypes combined due to the smaller number of events.

**Supplementary Fig. 5.**
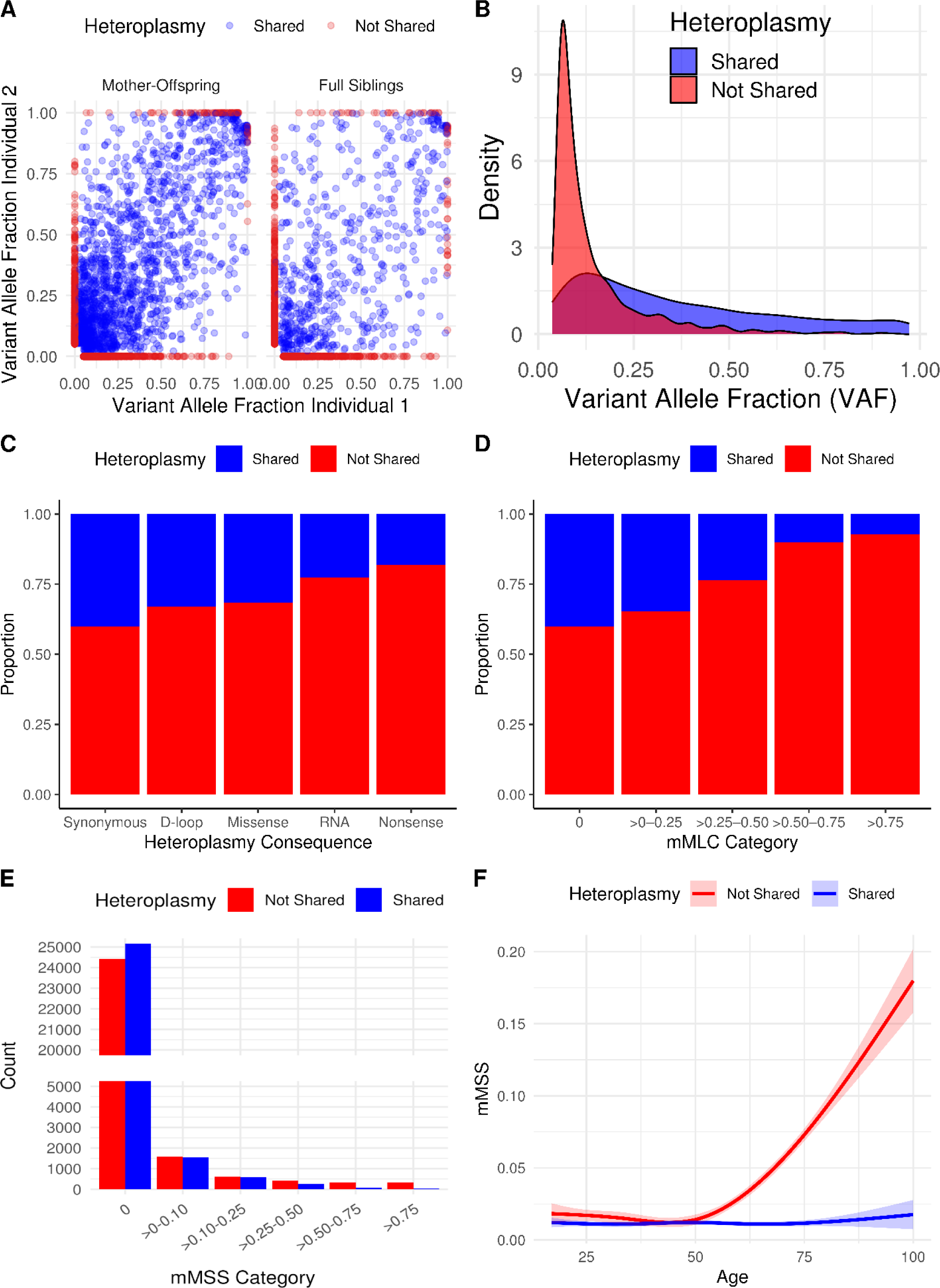
mtDNA heteroplasmy characteristsics stratified by sharing status in the All of Us cohort. **a,** Correlation of heteroplasmy VAFs stratified by relationship type. Left, Mother-Offspring have 1,676 shared variants, Pearson’s R=0.69; right, Full Siblings have 601 shared variants, Pearson’s R=0.56. **b,** Distribution of VAF. Shared heteroplasmy VAF is the average of the VAF for each relative pair. Not shared VAF is relative to the homoplasmy observed in the other relative. **c,** Proportion of heteroplasmies stratified by mutational consequence (synonymous, missense, nonsense) or for non-coding variation, complex (D-loop vs RNA). **d,** Proportion of heteroplasmies stratified by mMLC categories. **e,** mMSS count stratified by mMSS category. **f,** mMSS as a function of age adjusted for smoking status, sex, relative type, and top 4 mitochondrial PCs. Blue indicates shared mMSS and red indicates not shared mMSS.

**Supplementary Fig. 6.**
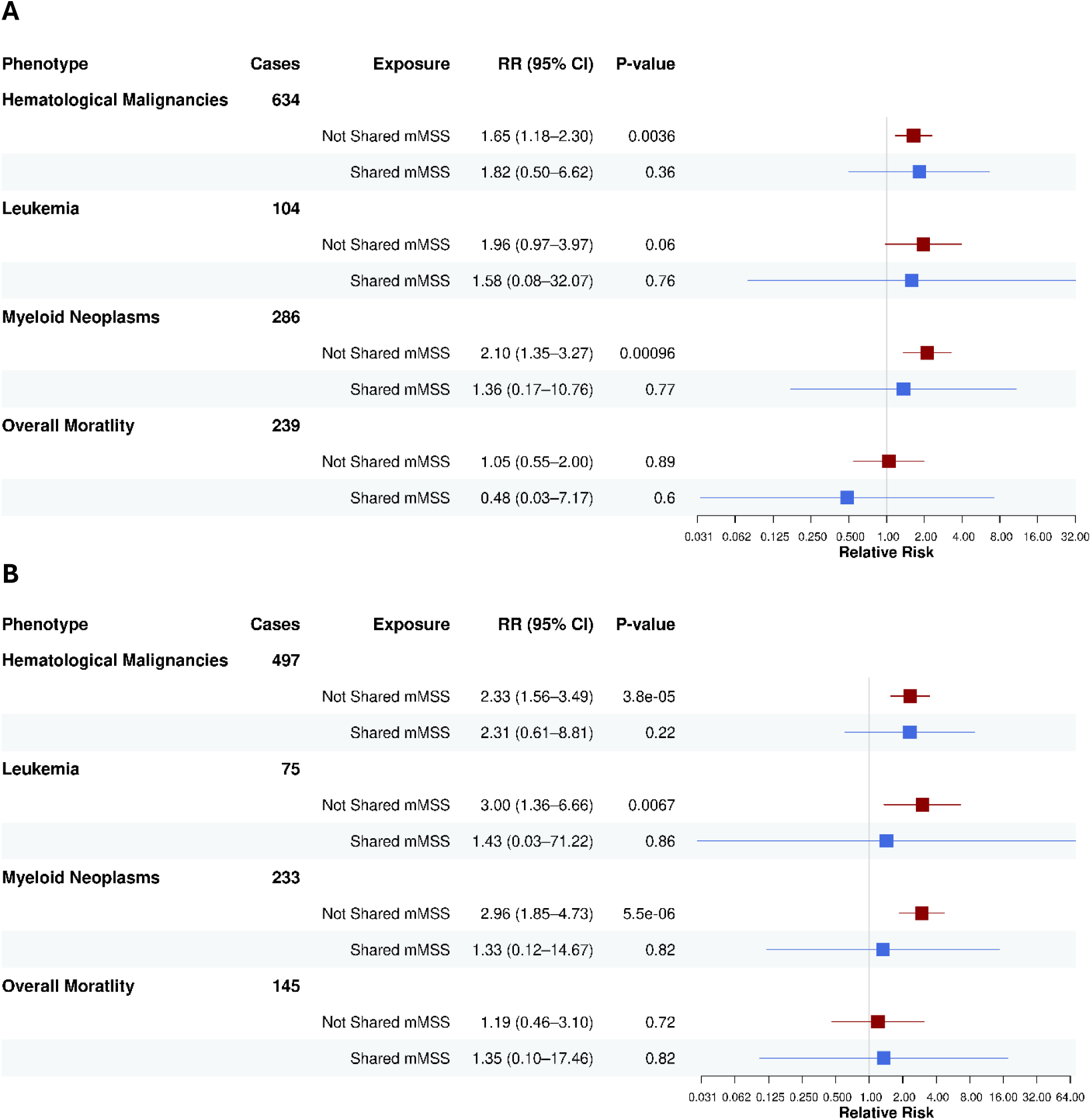
Forest plot depicting the association of shared and not shared mMSS with hematological malignancies and overall mortality in the All of Us Research Program. **a**, All ages. **b**, Age ≤ 70 years at study entry. Analyses are adjusted for age, sex, relative type, smoking exposure, and the top 4 mitochondrial PCs. Red indicates de novo mMSS and blue indicates shared mMSS. RR = relative risk; CI = confidence interval. Hematological malignancies excludes multiple myelomas, which we have previously shown are not associated with heteroplasmy.^5^

**Supplementary Fig. 7.**
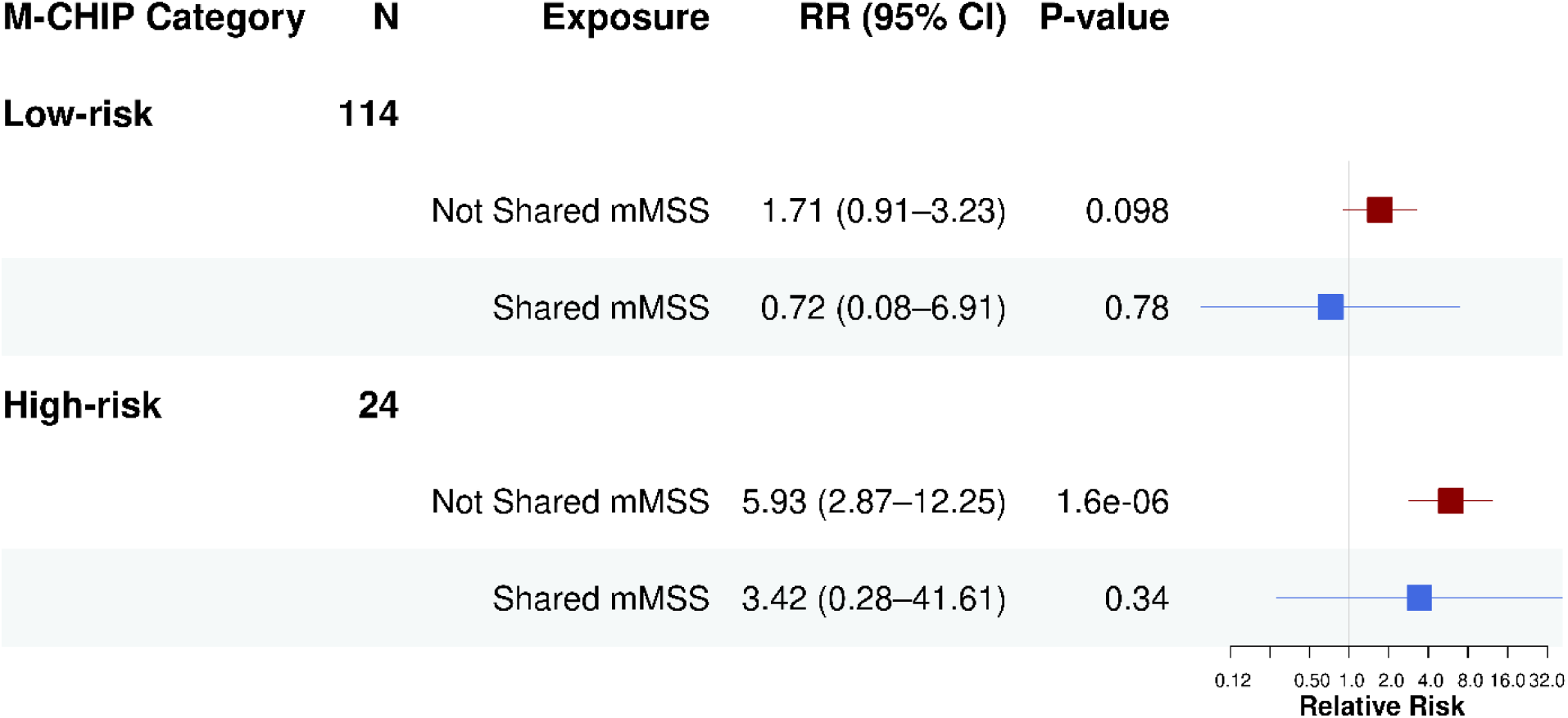
Forest plot depicting the association of shared and de novo mMSS with CHIP mutations in All of Us Research Program. Analyses are adjusted for age, sex, relative type, smoking exposure, and the top 4 mitochondrial PCs. Low-risk = single *DNMT3A* mutation; High-risk = *JAK2*, *SF3B1*, *SRSF2*, *ZRSR2*, *U2AF1*, *FLT3*, *IDH1*, *IDH2*, *NPM1*, *RUNX1*, *TP53*. Red indicates de novo mMSS and blue indicates shared mMSS. OR = odds ratio; CI = confidence interval.

**Supplementary Fig. 8.**
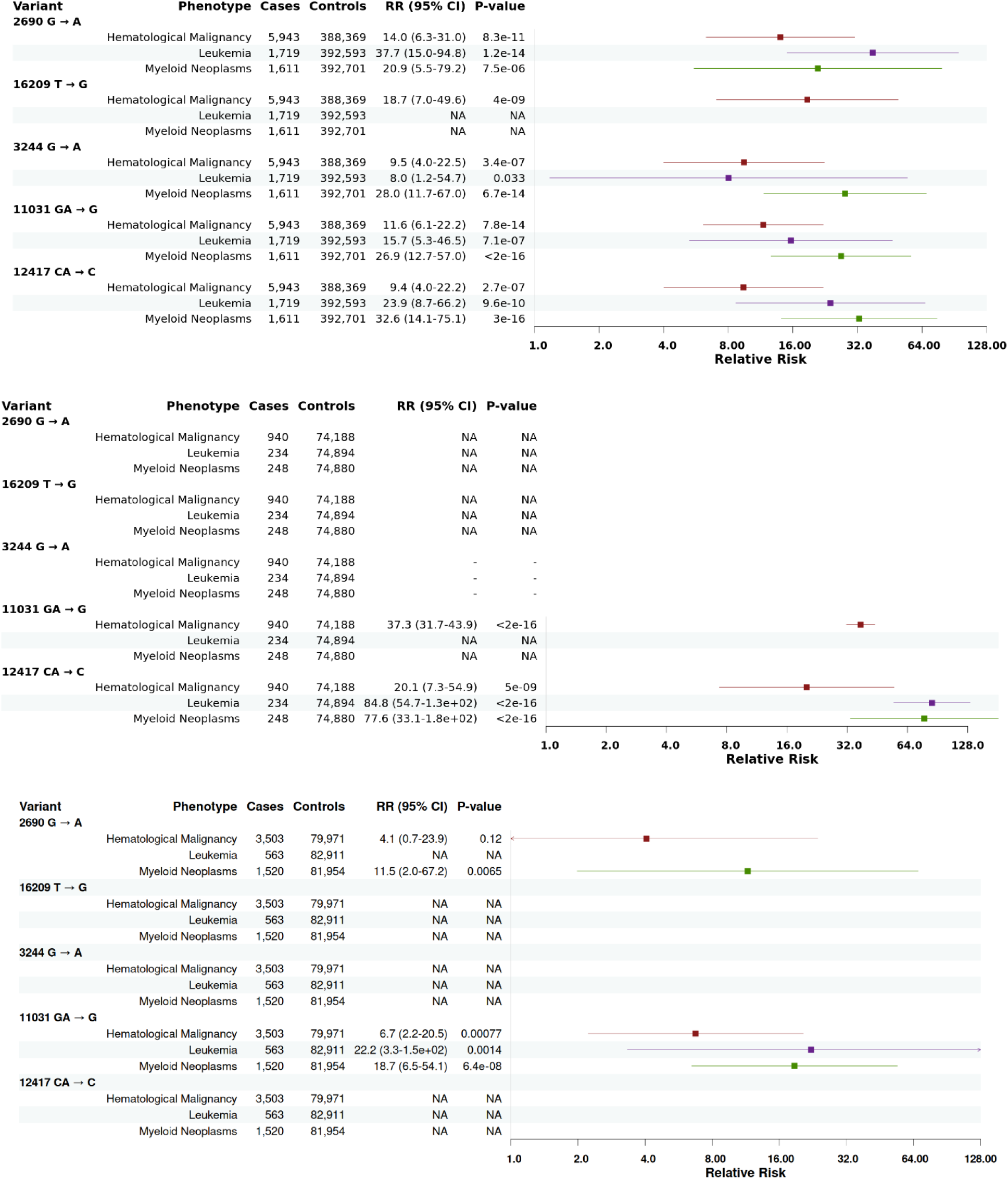
Forest plot depicting significant individual mtDNA variant associations with hematological malignancies. **a**, UKB white British ancestry, **b**, UKB non-white British ancestry, **c**, All of Us Research Program. Analyses were adjusted for age, sex, smoking exposure, mitochondrial continental haplogroup, and mtDNA PCs. “NA” denotes that no cases carried the variant, and “-” indicates that the variant was not present in the cohort.

**Supplementary Fig. 9.**
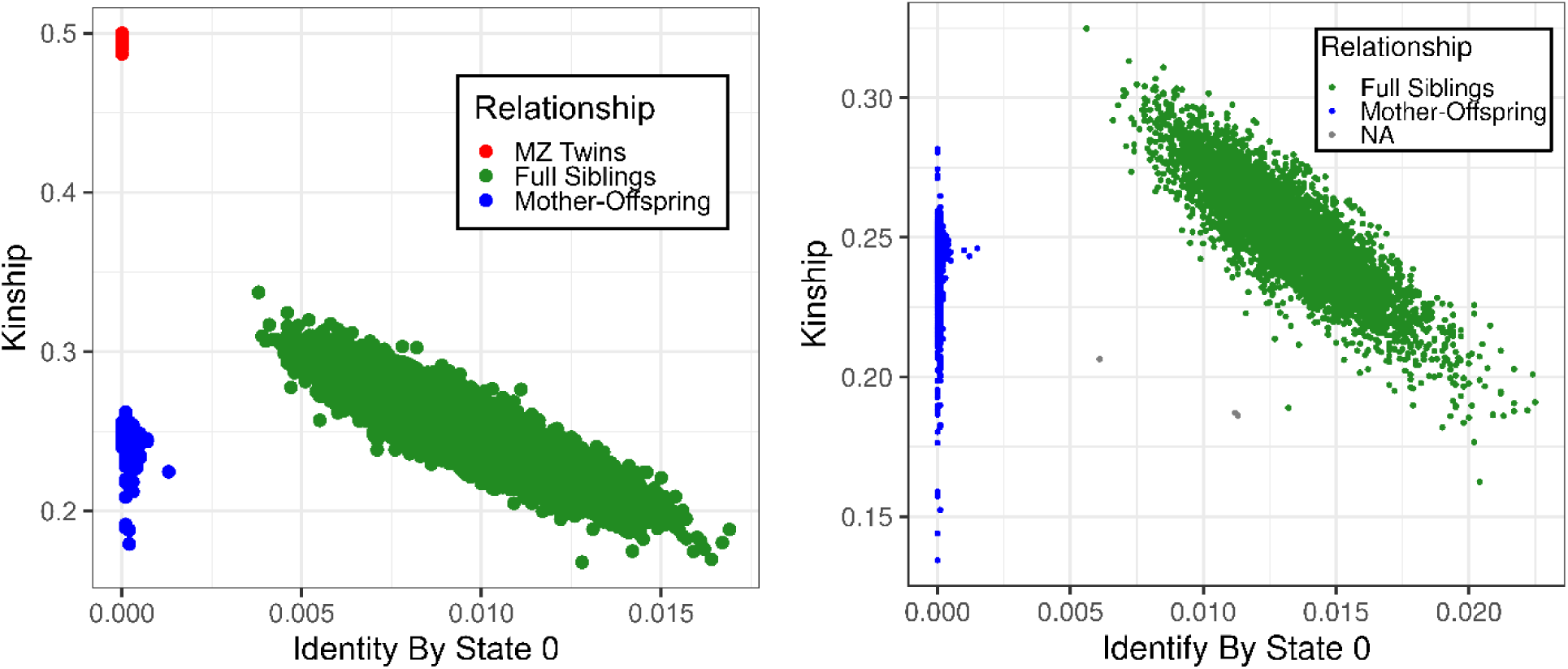
KING output for UKB (left) and All of Us (right). Relative pair types are readily distinguished by plotting Identity by State (IBS) 0 vs. the Kinship coefficient.

**Supplementary Fig. 10.**
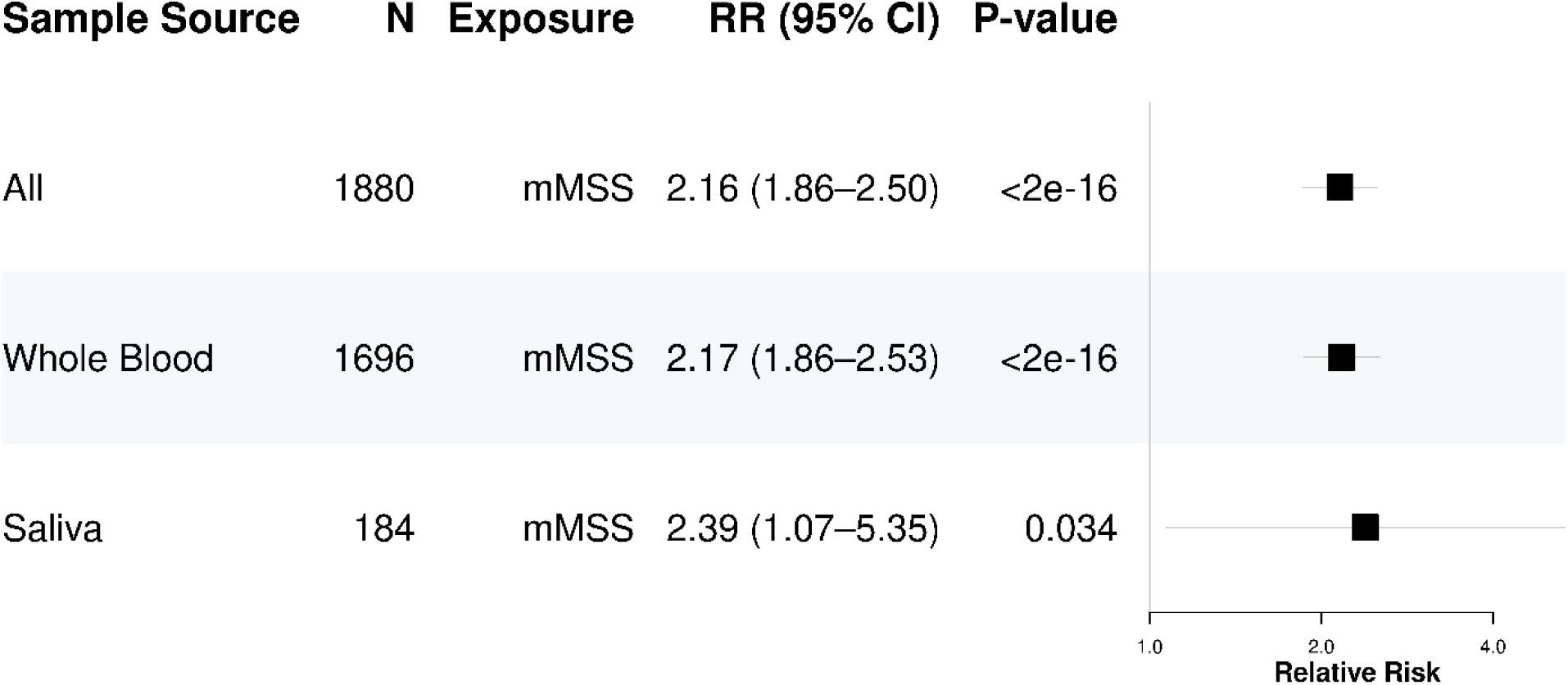
Comparison of association of mMSS with hematological malignancies by DNA source in a random cohort of All of Us Research Program. Analyses are adjusted for age, sex, smoking exposure, and the top 4 mitochondrial PCs. RR = relative risk; CI = confidence interval.

