## Supplemental Tables 1, 3, and 5 for "Deleterious mitochondrial heteroplasmy drives high-risk clonal hematopoiesis and hematological malignancy"

Supplementary Table 1. UKB Baseline Characteristics by Relationship Type

| **Variable** | **Full Sibling / MZ Twins (N=36148 / 340)** | **Mother (N=3910)** | **Offspring (N=3910)** | **Total (N=44308)** |
| --- | --- | --- | --- | --- |
| Age (years) |  |  |  |  |
| - Mean (SD) | 56.8 (7.3) | 66.0 (2.6) | 43.1 (2.3) | 56.4 (8.3) |
| - Median (Q1, Q3) | 58.0 (51.0, 62.0) | 66.0 (64.0, 68.0) | 43.0 (41.0, 45.0) | 58.0 (50.0, 63.0) |
| - Range | 40.0 - 70.0 | 56.0 - 70.0 | 39.0 - 53.0 | 39.0 - 70.0 |
| Sex |  |  |  |  |
| - Female | 21,135 (57.9%) | 3,910 (100.0%) | 2,374 (60.7%) | 27,419 (61.9%) |
| - Male | 15,353 (42.1%) | 0 (0.0%) | 1,536 (39.3%) | 16,889 (38.1%) |
| Self-identified Race |  |  |  |  |
| - Other | 1,120 (3.1%) | 114 (2.9%) | 153 (3.9%) | 1,387 (3.1%) |
| - White | 35,255 (96.9%) | 3,781 (97.1%) | 3,738 (96.1%) | 42,774 (96.9%) |
| - Missing | 113 | 15 | 19 | 147 |
| Mitochondrial Haplogroup |  |  |  |  |
| - EA | 35,381 (97.0%) | 3,784 (96.8%) | 3,784 (96.8%) | 42,949 (96.9%) |
| - AS | 631 (1.7%) | 67 (1.7%) | 67 (1.7%) | 765 (1.7%) |
| - AF | 476 (1.3%) | 59 (1.5%) | 59 (1.5%) | 594 (1.3%) |
| Smoking Status |  |  |  |  |
| - Never | 20,163 (55.5%) | 2,233 (57.5%) | 2,371 (60.9%) | 24,767 (56.1%) |
| - Previous | 12,533 (34.5%) | 1,406 (36.2%) | 984 (25.3%) | 14,923 (33.8%) |
| - Current | 3,635 (10.0%) | 244 (6.3%) | 540 (13.9%) | 4,419 (10.0%) |
| - Missing | 157 | 27 | 15 | 199 |
| Smoking Frequency |  |  |  |  |
| - All_days | 11,475 (31.5%) | 1,141 (29.2%) | 1,029 (26.4%) | 13,645 (30.8%) |
| - Never | 20,161 (55.3%) | 2,233 (57.2%) | 2,371 (60.7%) | 24,765 (55.9%) |
| - Occasional | 4,690 (12.9%) | 511 (13.1%) | 495 (12.7%) | 5,696 (12.9%) |
| - Unclear | 140 (0.4%) | 21 (0.5%) | 9 (0.2%) | 170 (0.4%) |
| - Missing | 22 | 4 | 6 | 32 |
| Smoking Duration (years) |  |  |  |  |
| - Mean (SD) | 8.1 (14.0) | 9.2 (16.0) | 5.3 (9.8) | 8.0 (13.9) |
| - Median (Q1, Q3) | 0.0 (0.0, 13.0) | 0.0 (0.0, 15.0) | 0.0 (0.0, 6.0) | 0.0 (0.0, 13.0) |
| - Range | 0.0 - 58.0 | 0.0 - 58.0 | 0.0 - 39.0 | 0.0 - 58.0 |
| - Missing | 22 | 4 | 6 | 32 |
| Mortality |  |  |  |  |
| - No | 33,615 (92.1%) | 3,322 (85.0%) | 3,822 (97.7%) | 40,759 (92.0%) |
| - Yes | 2,873 (7.9%) | 588 (15.0%) | 88 (2.3%) | 3,549 (8.0%) |
| Hematologic Malignancy |  |  |  |  |
| - No | 35,929 (98.5%) | 3,829 (97.9%) | 3,882 (99.3%) | 43,640 (98.5%) |
| - Yes | 559 (1.5%) | 81 (2.1%) | 28 (0.7%) | 668 (1.5%) |
| Leukemia |  |  |  |  |
| - No | 36,313 (99.5%) | 3,890 (99.5%) | 3,905 (99.9%) | 44,108 (99.5%) |
| - Yes | 175 (0.5%) | 20 (0.5%) | 5 (0.1%) | 200 (0.5%) |
| Myeloid Neoplasm |  |  |  |  |
| - No | 36,346 (99.6%) | 3,890 (99.5%) | ≥3,905 (99.9%) | 44,142 (99.6%) |
| - Yes | 142 (0.4%) | 20 (0.5%) | ≤5 (0.1%) | 166 (0.4%) |
| M-CHIP |  |  |  |  |
| - No | 31,949 (92.2%) | 3,306 (88.9%) | 3,584 (95.5%) | 38,839 (92.2%) |
| - Yes | 2,718 (7.8%) | 414 (11.1%) | 167 (4.5%) | 3,299 (7.8%) |
| - Missing | 1,821 | 190 | 159 | 2,170 |
| Data are presented as mean (SD) or n (%). Hematological Malignancies excludes multiple myeloma, which we have previously shown is not associated with heteroplasmy.(REF). EA = European Ancestry; AS = Asian Ancestry; AF = African Ancestry; M-CHIP = myeloid CHIP. MZ Twins are combined with Full Siblings to avoid small cell counts and possible reidentification. | | | | |

Supplementary Table 3. All of Us Baseline Characteristics by Relationship Type

| **Variable** | **Full Sibling (N=9436)** | **Mother (N=9113)** | **Offspring (N=9113)** | **Total (N=27662)** |
| --- | --- | --- | --- | --- |
| Age (years) |  |  |  |  |
| - Mean (SD) | 52.7 (16.2) | 59.3 (11.7) | 33.8 (11.7) | 48.7 (17.2) |
| - Median (Q1, Q3) | 56.0 (41.0, 65.0) | 58.0 (51.0, 67.0) | 32.0 (24.0, 41.0) | 50.0 (34.0, 62.0) |
| - Range | 17.0 - 97.0 | 33.0 - 101.0 | 17.0 - 76.0 | 17.0 - 101.0 |
| Self-identified Race |  |  |  |  |
| - Black | 1,988 (21.1%) | 2,022 (22.2%) | 2,052 (22.5%) | 6,062 (21.9%) |
| - Other / Not Indicated | 3,103 (32.9%) | 3,584 (39.3%) | 3,590 (39.4%) | 10,277 (37.2%) |
| - White | 4,345 (46.0%) | 3,507 (38.5%) | 3,471 (38.1%) | 11,323 (40.9%) |
| Mitochondrial Haplogroup |  |  |  |  |
| - AS | 2,224 (23.6%) | 2,516 (27.6%) | 2,516 (27.6%) | 7,256 (26.2%) |
| - EA | 4,810 (51.0%) | 4,025 (44.2%) | 4,025 (44.2%) | 12,860 (46.5%) |
| - AF | 2,402 (25.5%) | 2,572 (28.2%) | 2,572 (28.2%) | 7,546 (27.3%) |
| Smoking Status |  |  |  |  |
| - 100 Cigs Lifetime: No | 5,816 (61.6%) | 5,885 (64.6%) | 6,534 (71.7%) | 18,235 (65.9%) |
| - 100 Cigs Lifetime: Yes | 3,620 (38.4%) | 3,228 (35.4%) | 2,579 (28.3%) | 9,427 (34.1%) |
| Sex |  |  |  |  |
| - Female | 6,341 (67.2%) | 9,113 (100.0%) | 6,633 (72.8%) | 22,087 (79.8%) |
| - Male | 3,095 (32.8%) | 0 (0.0%) | 2,480 (27.2%) | 5,575 (20.2%) |
| Mortality |  |  |  |  |
| - No | 7,272 (98.7%) | 6,955 (98.4%) | 6,941 (99.5%) | 21,168 (98.9%) |
| - Yes | 93 (1.3%) | 111 (1.6%) | 35 (0.5%) | 239 (1.1%) |
| - Missing | 2,071 | 2,047 | 2,137 | 6,255 |
| Hematologic Malignancy |  |  |  |  |
| - No | 7,119 (96.7%) | 6,828 (96.6%) | 6,826 (97.8%) | 20,773 (97.0%) |
| - Yes | 246 (3.3%) | 238 (3.4%) | 150 (2.2%) | 634 (3.0%) |
| - Missing | 2,071 | 2,047 | 2,137 | 6,255 |
| Leukemia |  |  |  |  |
| - No | 7,310 (99.3%) | 7,033 (99.5%) | ≥6,940 (99.7%) | 21,303 (99.5%) |
| - Yes | 55 (0.7%) | 33 (0.5%) | ≤21 (0.3%) | 104 (0.5%) |
| - Missing | 2,071 | 2,047 | 2,137 | 6,255 |
| Myeloid Neoplasm |  |  |  |  |
| - No | 7,256 (98.5%) | 6,963 (98.5%) | 6,902 (98.9%) | 21,121 (98.7%) |
| - Yes | 109 (1.5%) | 103 (1.5%) | 74 (1.1%) | 286 (1.3%) |
| - Missing | 2,071 | 2,047 | 2,137 | 6,255 |
| M-CHIP |  |  |  |  |
| - No | 9,213 (97.6%) | 8,851 (97.1%) | 9,006 (98.8%) | 27,070 (97.9%) |
| - Yes | 223 (2.4%) | 262 (2.9%) | 107 (1.2%) | 592 (2.1%) |
| Data are presented as mean (SD) or n (%). Hematological Malignancies excludes multiple myeloma, which we have previously shown is not associated with heteroplasmy.(REF). EA = European Ancestry; AS = Asian Ancestry; AF = African Ancestry; M-CHIP = myeloid CHIP. Cell counts <21 are not reported to avoid potential reidentification. | | | | |

**Supplementary Table 5. Variant annotations for significant variants identified via REGENIE in the UKB white British Population.**

| **Variant** | **Position** | **Gene** | **Complex** | **Mutation Consequence** | **mMLC** | **MAF in UKB** | **VAF Range** |
| --- | --- | --- | --- | --- | --- | --- | --- |
| 2690 G → A | 2690 | RNR2 | RRNA | Non-Coding Transcript  Exon Variant | 0.768 | 4.1e-05 | (0.051 - 0.151) |
| 3244 G → A | 3244 | TRNL1 | TRNA | Non-Coding Transcript  Exon Variant | 0.848 | 3.5e-05 | (0.057 - 0.185) |
| 11031 GA → G | 11031 | ND4 | Complex I | Frameshift | - | 5.7e-05 | (0.052 - 0.226) |
| 12417 CA → C | 12417 | ND5 | Complex I | Frameshift | - | 5.7e-05 | (0.051 - 0.210) |
| 16209 T → G | 16209 | DLOOP | DLOOP | Intergenic | 0.015 | 3.3e-05 | (0.086 - 1.000) |

mMLC is the modified mitochondrial DNA local constraint score. Indels do not have an assigned mMLC score. MAF was calculated as the total number of carriers of the mutations at any VAF ≥ 0.05 divided by the total number of individuals present in the genetic data file. MAF is presented as a decimal and not as a percentage.
